# The complex interplay between risk tolerance and the spread of infectious diseases

**DOI:** 10.1101/2024.07.01.24309771

**Authors:** Maximilian Nguyen, Ari Freedman, Matthew Cheung, Chadi Saad-Roy, Baltazar Espinoza, Bryan Grenfell, Simon Levin

**Affiliations:** Lewis-Sigler Institute, Princeton University, Princeton, NJ 08544; Department of Ecology and Evolutionary Biology, Princeton University, Princeton, NJ, USA; Program in Applied and Computational Mathematics, Princeton University, Princeton, NJ, USA; Miller Institute for Basic Research in Science, Department of Integrative Biology, University of California, Berkeley, Berkeley, CA, USA; Biocomplexity Institute, University of Virginia, Charlottesville, VA, USA

## Abstract

Risk-driven behavior provides a feedback mechanism through which individuals both shape and are collectively affected by an epidemic. We introduce a general and flexible compartmental model to study the effect of heterogeneity in the population with regards to risk tolerance. The interplay between behavior and epidemiology leads to a rich set of possible epidemic dynamics. Depending on the behavioral composition of the population, we find that increasing heterogeneity in risk tolerance can either increase or decrease the epidemic size. We find that multiple waves of infection can arise due to the interplay between transmission and behavior, even without the replenishment of susceptibles. We find that increasing protective mechanisms such as the effectiveness of interventions, the number of risk-averse people in the population, and the duration of intervention usage reduces the epidemic overshoot. When the protection is pushed past a critical threshold, the epidemic dynamics enter an underdamped regime where the epidemic size exactly equals the herd immunity threshold and overshoot is eliminated. Lastly, we can find regimes where epidemic size does not monotonically decrease with a population that becomes increasingly risk-averse.

## 1 Introduction

Recent outbreaks such as the COVID-19 pandemic, the 2014 Ebola outbreak, the 2009 influenza A (H1N1) pandemic, and the 2002 SARS epidemic brought to light many of the challenges of mounting an effective and unified epidemic response in a country as large and as diverse as the United States. Particularly during the COVID-19 pandemic, people were split in opinion on questions such as the origin of the virus [1], whether they would social distance or wear a mask [2–4], or whether the country should even have a pandemic response at all [5]. As time progressed, the situation became more dire and the death toll accumulated. People then had a new battery of questions to address, such as whether or not they would adhere to mandatory lock-downs [6–8] or whether they felt comfortable using the new mRNA vaccines [9, 10]. Compounding the issue were the multiple streams of information and potential misinformation spread through social media and other channels [11–14]. People’s stances on the questions and issues were diverse, arising from the milieu of differences in culture, geography, scientific education, sources of information, political leanings, and individual identity [15–17].

Taken altogether, these differences within the population reflect a spectrum of people’s risk tolerances to a circulating infectious disease. For any given intervention, such as social distancing, wearing a mask, or taking a vaccine, each person in the population falls somewhere on a spectrum of willingness to adopt the intervention. Given a threat level of an infectious disease in the population, some people will readily wear masks, whereas other people will refuse to.

In this study, we aim to analyze the impact of heterogeneity in risk tolerance and the resulting behavioral response on the dynamics of epidemics. We seek to add to a burgeoning literature on the impact of human behavior in epidemic response [18–32], which the recent pandemic highlighted as an area for further exploration in preparation for the next large scale global health crisis [33–35]. To study the impact of heterogeneity in risk tolerance on epidemic dynamics, we introduce a simple and flexible modeling framework based on ordinary differential equations that can be used for different interventions and an arbitrary partitioning of the population with regard to risk tolerance and behavioral responses. We will examine and discuss potential interesting outcomes that can arise from coupling individual-level preferences and population-level epidemiology.

## 2 Results

### Model of Adaptive Intervention Usage under Heterogeneous Risk Tolerance

Here we assume people’s risk aversion manifests as the rate at which they adopt individual interventions in response to an infectious disease outbreak. The intuition underlying this paradigm is that more risk-averse individuals are more sensitive to becoming sick and thus will adopt interventions at a faster rate than more risk-tolerant people. We consider the following SPIR compartmental model of a population with *n* differing levels of risk tolerance (1-4). This model features four types of classes: unprotected susceptible (*S*), protected susceptible (*P*), infectious (*I*), and recovered with permanent immunity (*R*). Since there are *n* differing levels of risk tolerance, we subdivide the susceptible population into *n* discrete groups indexed by *i*, where *i* ∈{1, 2, …, *n*}. Each tolerance level is characterized by an intervention adoption rate parameter (*λ*_*i*_) and an intervention relaxation rate parameter (*δ*_*i*_). Transitions of susceptibles between their unprotected class (*S*_*i*_) and their corresponding protected class (*P*_*i*_) are governed by the corresponding parameters of the same index (*λ*_*i*_, *δ*_*i*_). Overall, the system is governed by 3 + 2*n* parameters: a transmission rate parameter (*β*), a recovery rate parameter (*γ*), an intervention effectiveness parameter (*ϵ*), and an intervention adoption rate (*λ*_*i*_) and intervention relaxation rate (*δ*_*i*_) for each tolerance level. Since we always assume there are no protected individuals initially, the dynamics of the infected compartment is initially only a function of the transmission and recovery parameters. Thus we can use the typical definition for the basic reproduction number 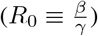.

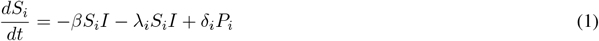

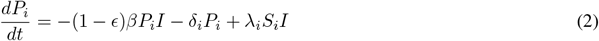

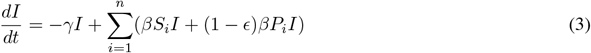

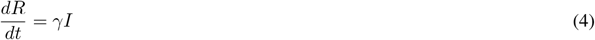

The transition from the unprotected susceptible state to the protected susceptible state represents individuals implementing an intervention that confers them protection against disease transmission from an infected individual. The rate at which intervention adoption occurs may be driven by individuals considering information such as the epidemic incidence rate (e.g. cases per day), the total number of infected individuals in the population (e.g. total number of active cases), and mortality rate (e.g. deaths per day) [22]. Here we assume that individuals have knowledge about the total number of infected individuals (*I*) and respond accordingly. Parameterizing each person’s individual risk tolerance by *λ*_*i*_, we assume each individual person adopts an intervention at a rate *λ*_*i*_*I*. Then, if there are *S*_*i*_ number of people that behave exactly the same (i.e. have the same level of risk-aversion), then at the population scale there is a collective adoption rate of *λ*_*i*_*S*_*i*_*I*. The same reasoning holds for each of the *n* tolerance levels. We also consider a model where the adoption rate is driven by individuals reacting to the incidence rate (Supplemental Materials); while this produces a more complex mathematical model, the results are qualitatively similar.

The effectiveness of the intervention being used is captured by the parameter *ϵ*, which linearly scales down the transmission rate between infected and protected susceptibles. In the limit of *ϵ* = 1, the intervention is perfectly effective and protected individuals cannot become infected. In the limit of *ϵ* = 0 then the intervention is completely ineffective, which reduces the model to an SIR model without interventions. For simplicity, we assume each epidemic features only a single type of intervention (whether that be masking, social distancing, vaccines, etc.) and that the effectiveness of an intervention is identical across the population. In reality, multiple interventions may be available concurrently, which would drive additional variation in behavior due to differences in risk sensitivity across the population.

This model allows for protected individuals to relax their usage of interventions, becoming unprotected in the process. Here, individuals in the protected class can relax back to the unprotected class through an infection-independent rate (*δ*_*i*_*P*_*i*_) that is governed by the intervention relaxation rate parameter (*δ*_*i*_) for each tolerance level. In the limit of *δ*_*i*_ = 0, an intervention is irreversible, which would represent an intervention such as vaccines with permanent immunity. When *δ*_*i*_ is non-zero, individuals are using interventions such as masking or social distancing. This relaxation rate is motivated by factors such as psychological fatigue of social distancing [36, 37] and physical discomfort with wearing masks [38]. In general, we will consider the regime where the relaxation rate *δ* is of comparable scale or smaller than the transmission scale (i.e. *δ* ≤ *β*). This reflects intuition that people are likely to continue to protect themselves with interventions even beyond an initial outbreak [39].

For simplicity, we consider the model for the case when *n* = 1 and *n* = 2. A schematic for these two cases is shown in Figure 1. However, the framework is general and can be extended to any discrete number of groups.

**Figure 1:**
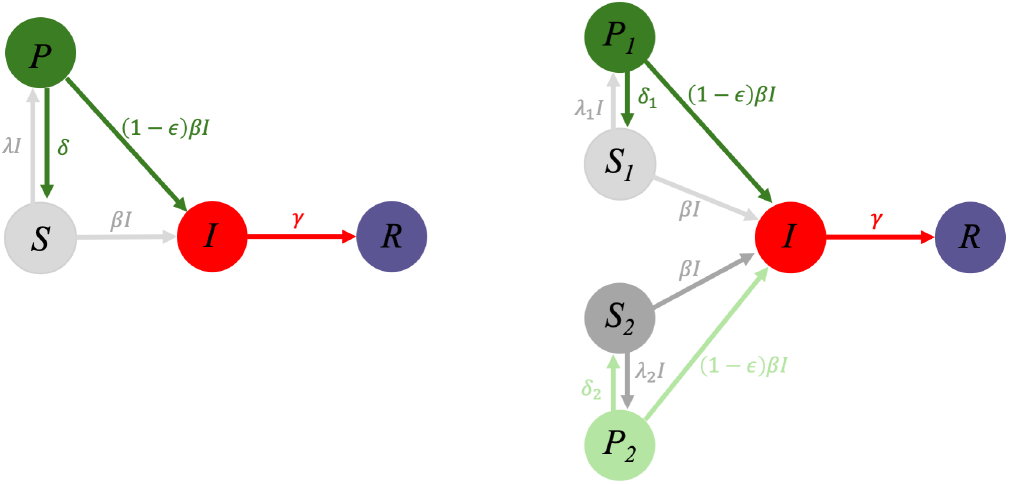
Flow diagram for an SIR model with adaptive interventions for either (a) a population with homogeneous risk tolerance or (b) a heterogeneous population with two different levels of risk tolerance. Susceptible individuals can access a more protected susceptible state through usage of interventions. The transition rate to the protected state depends on the incidence level. The protected state offers a 1 − *ϵ* reduction in transmission rate over the normal susceptible state.

For convention, when there are two susceptible classes, we assume the first susceptible class (*S*_1_) has a lower risk tolerance for becoming infected (i.e. more risk-averse). As a result, these individuals more readily adopt the intervention (i.e. *λ*_1_ > *λ*_2_), making individuals in this class transition more rapidly to the protected susceptible state (*P*_1_). The second susceptible class (*S*_2_) is more risk tolerant (i.e. more risk-taking), and thus is less eager to use the intervention, making individuals in this class transition more slowly to their protected susceptible state (*P*_2_).

The model introduced here has several similarities and distinctions compared to existing behavior-disease models in the literature[19, 22, 28, 40–42], particularly to the global, prevalence-based spread of fear of disease of Perra et al. [43]. Our model shares the spirit of a disease contagion occuring at the same time as a social contagion. While the Perra model couches the social contagion in the language of individuals and fear, here we take the viewpoint that individuals have a personal characteristic called risk-tolerance, that encapsulates fear and other underlying biopsychosocial factors. The main feature of our model is the consideration of heterogeneity in the population, allowing for the population to be split into arbitrary factions of risk-tolerance. In addition, compared to the Perra model, the model here allows for people to remove protection at a rate independent of the disease.

### Adaptive Adoption of Interventions Can Produce Damped Oscillations

The coupling of intervention usage to the incidence rate and the resulting adaptive changes enables the epidemic dynamics to display a much richer set of behavior over the simple SIR model. From Figure 2, we see this particular set of conditions can deterministically produce multiple waves of infection, even when vital dynamics (i.e. birth and death processes) are not considered.

**Figure 2:**
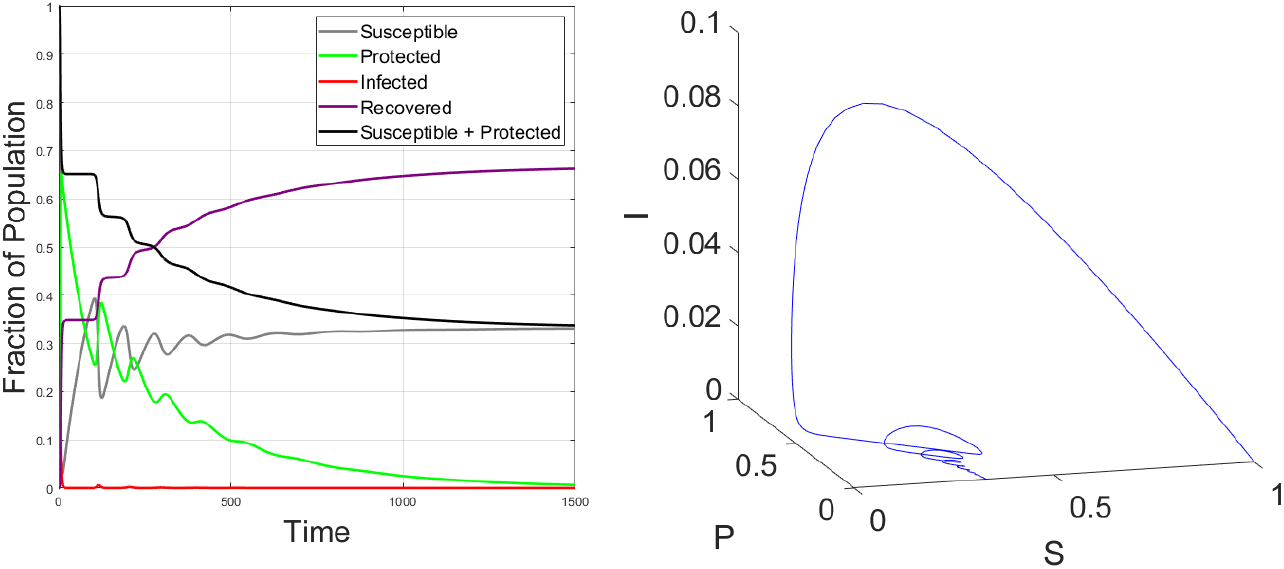
Time series for population with homogeneous risk tolerance and adaptive intervention usage and the corresponding phase space trajectory indicate the possibility for damped oscillations. Parameter values: *β* = 3, *γ* = 1, *ϵ* = 0.8, *λ* = 10, *δ* = 0.01. Initial conditions: *I*(0) = 10^*−*^6, *S*(0) = 1 − *I*(0), *P* (0) = *R*(0) = 0.

Evidence for cycling of individuals between using interventions and not using interventions during the COVID pandemic can be seen in longitudinal usage [44–47] data. The possibility for these oscillations highlight the intimate connection between individual human behavior and intervention usage in shaping the dynamics of epidemics, while also be affected by the collective decision of everyone in the population. The coupling of behavior and epidemiology here provides a feedback mechanism where an increasing incidence rate prompts more individuals to adopt an intervention, which lowers the overall incidence rate; however, as the epidemic wanes and factors such as fatigue or discomfort set in, people begin dropping their usage of interventions, which may eventually lead to another wave of outbreaks if enough people become unprotected while infected individuals still remain, and then the cycle can be repeated. The oscillatory phenomenology here is reminiscent of several other behavioral models in the literature [48, 49].

### Protective Mechanisms Saturate in Underdamped Regime that Eliminates Epidemic Overshoot

One might have the intuition that having more people that will more readily adopt an intervention (i.e. mask, social distance, or vaccinate) or increasing the effectiveness of the intervention in reducing transmission will further decrease the size of the epidemic. Here we define epidemic size to be the cumulative number of infections over the course of the epidemic, or equivalently, the total depletion of susceptible people over the course of the epidemic. While we find this intuition to be mostly correct, we unexpectedly find that the protection conferred by either of these mechanisms can saturate once a critical parameter threshold has been passed.

In Figure 3, *left*, we see that increasing the effectiveness of the intervention or increasing the fraction of the population that are risk-averse monotonically decreases the epidemic size. However, in the dark blue region (which we will refer to as the underdamped regime) where both protection mechanisms are at their highest, we see no further reduction in the epidemic size. This regime corresponds to an epidemic where the epidemic size exactly equals the herd immunity threshold, which is the minimum number of susceptibles that must be eliminated from the pool to prevent further outbreak from occurring. Since the epidemic size is the sum of the herd immunity threshold and the epidemic overshoot, this further implies that epidemic overshoot is eliminated in the underdamped regime.

**Figure 3:**
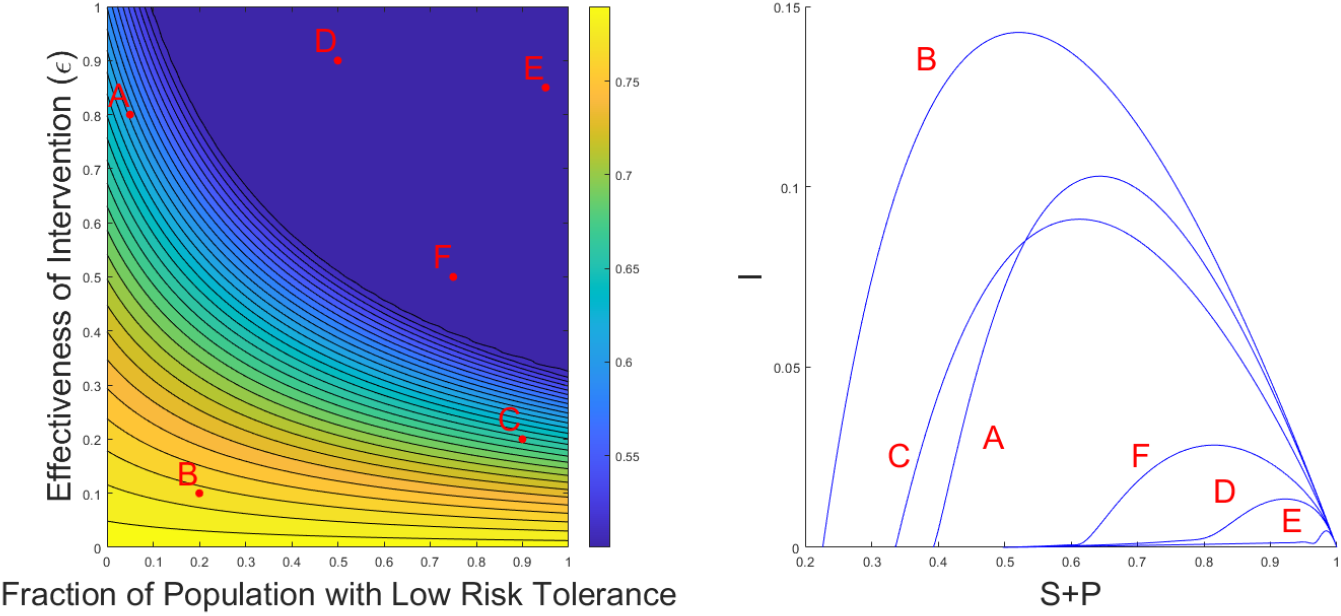
Left. Epidemic size as a function of varying the fraction of the population that are low-risk tolerance (i.e. those with higher *λ*). Right. Corresponding orbits in the I versus S+P plane for the sampled points in parameter space. Parameter values: *β* = 2, *γ* = 1, *λ*_1_ = 100, *δ*_1_ = 0.1, *λ*_2_ = 1, *δ*_2_ = 0.1,. Initial conditions: *I*(0) = 10^*−*^8, *P*_1_(0) = *P*_2_(0) = *R*(0) = 0.

The orbits of the dynamics from different areas of this parameter space are shown in Figure 3, *right*. We notice that orbits in the overdamped region (Orbits A, B, C) are parabolas and that each of those orbits ends at a different final number of susceptibles. In contrast, orbits in the underdamped regime (Orbits D, E, F) are qualitatively different in that they display long tails that converge to the same final number of susceptibles. We see that even though the final epidemic size is the same throughout the underdamped region, the trajectories to reach the same final epidemic size can look qualitatively different.

Figures S4-S5 show a larger sampling of trajectories if one fixes either the fraction of the population with low risk tolerance or the intervention effectiveness respectively. It becomes clear that at the border of the underdamped region, we can see a clear change in the qualitative behavior of the trajectories as the threshold is crossed. Under some assumptions, one can prove that that the epidemic overshoot is eliminated in the underdamped regime (Supplemental Information).

However, we should make the point that this is not evidence that highly effective interventions are a waste or that the overall population should tolerate risky behavior. As this is a model with a large parameter space, we cannot visualize all of it. If we could, we would find many parameter regimes where the protection mechanisms never reach a critical threshold, which implies the conventional intuition of increasing intervention effectiveness and having more risk-averse people always being beneficial applies.

### The Threshold to the Underdamped Regime is Reduced when Intervention Usage is Prolonged

The transition to the underdamped regime is more easily accessed when the usage of interventions is prolonged (or equivalently when the rate at which protected individuals relax back into the regular susceptibility classes decreases). Consider the following scenario which is identical to the previous setup, except now the intervention reversion rate (*δP*_*i*_) has been reduced by an order of magnitude (Figure 4). This corresponds to a scenario where people continue to use the intervention (i.e. such as wearing masks or social distancing) on a timescale significantly longer than the transmission timescale 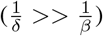.

**Figure 4:**
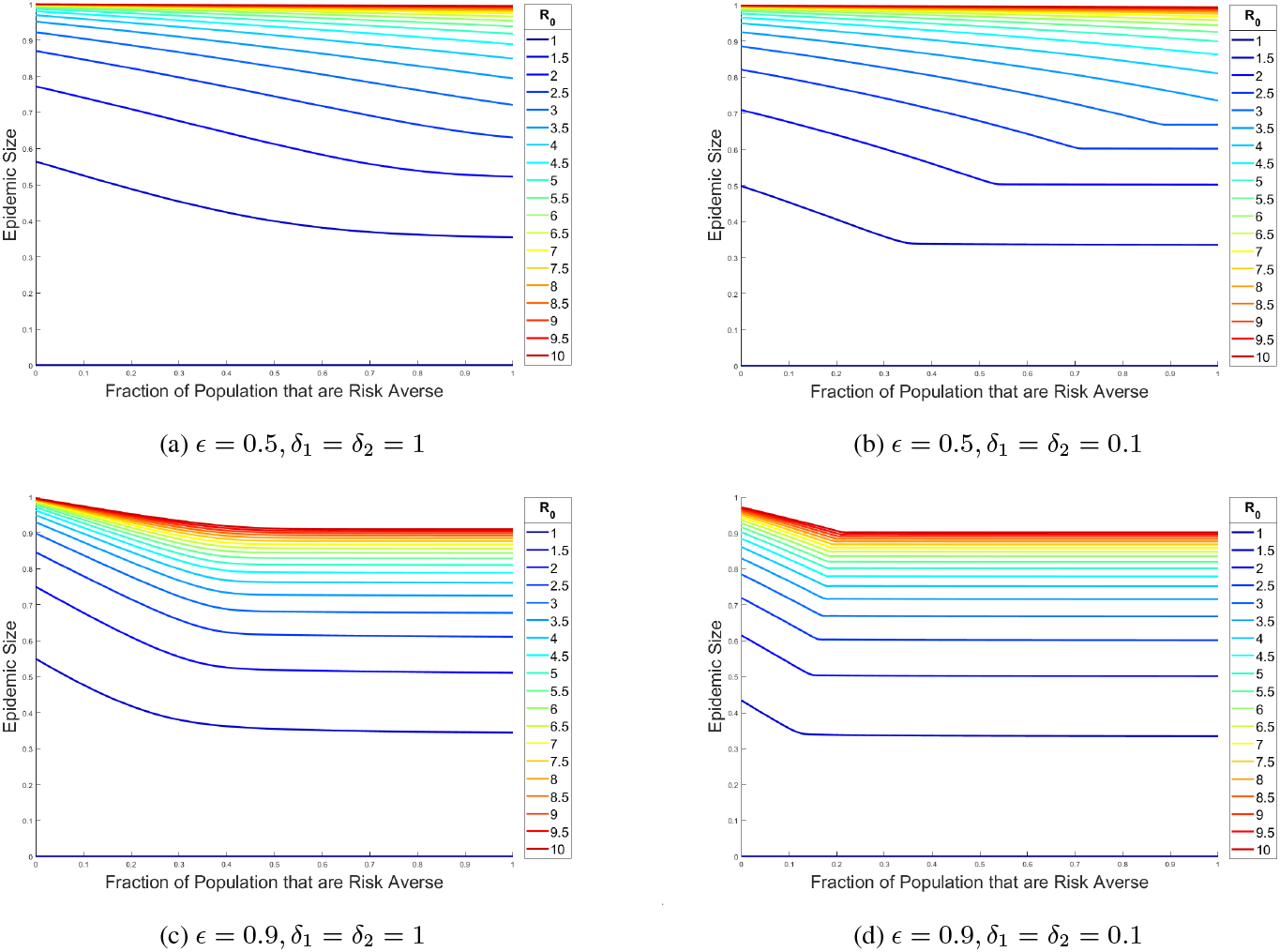
Final epidemic size versus fraction of population that are risk-averse (*S*_1_). Simulations in the left column have a higher *δ* than simulations in the right column. Parameter values: *R*_0_ = *β, γ* = 1, *λ*_1_ = 100, *λ*_2_ = 1. Initial conditions: *I*(0) = 10^*−*^8, *P*_1_(0) = *P*_2_(0) = *R*(0) = 0.

This suggests that increasing the timescale at which individuals continue to use interventions decreases the number of risk-averse individuals needed to achieve the same epidemic size. This is reflected in the horizontal shift of the transition region to the left when comparing Figures in the left column and the right column (Figure 4). We also briefly investigate what happens if we allow for heterogeneous relaxation rates between the difference groups, which is more reflective of what occurs in reality, finding the epidemic size to be more attune to the slower relaxation time scale (Supplemental Information).

### Increasing Heterogeneity in Risk Tolerance can Either Increase or Decrease the Epidemic Size

There are many types of heterogeneity that may be present in the population, arising from differences in characteristics such as biological, cultural factors, and socioeconomic factors. The effect of each type of heterogeneity on disease dynamics is an area of active study. Some previous literature has suggested that increasing heterogeneity in the population through increasing the variation in their contact patterns, age, or general susceptibility results in a either a reduction or no change in the epidemic size [50–53].

We find in this model of heterogeneous behavior that it is possible to switch from a regime where increasing the heterogeneity in risk tolerance results in a decrease in epidemic size to a regime where increasing the heterogeneity in risk tolerance results in an increase in epidemic size. This result also does not have to necessarily be due to a dramatic shift in parameters. From Figure 5, we see this shift can arise from solely varying the fraction of the population with low risk tolerance by a small amount.

**Figure 5:**
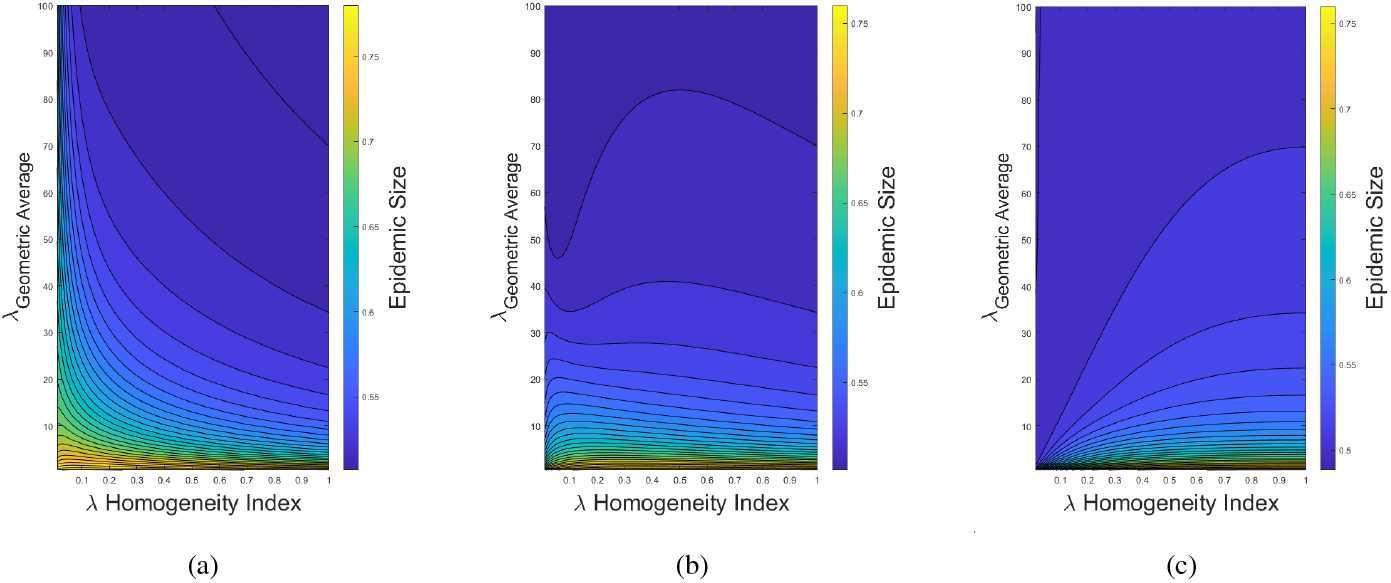
Epidemic size under differing levels of heterogeneity in the adoption rate for interventions. The mean adoption rate of the two groups (i.e. geometric average of *λ*_1_, *λ*_2_) is compared to the difference in the two adoption rates as parameterized by a homogeneity index (see Methods for definition). *Left* is when the fraction of the population with low risk tolerance (*x*_1_) is 0.2, *center* is when *x*_1_ = 0.35, *right* is when *x*_1_ = 0.5. Parameter values: *β* = 2, *γ* = 1, *ϵ* = 0.7, *δ*_1_ = *δ*_2_ = 0.5. Initial conditions: *I*(0) = 10^*−*^8, *P*_1_(0) = *P*_2_(0) = *R*(0) = 0.

The intuition underlying this result is that when the average adoption rate (*λ*_*average*_), which is expressed as a (geometric or arithmetic) weighted mean of the adoption rates of the two groups, is fixed at a constant level, then the epidemic size can be suppressed through either varying the fraction of the population in each group or through varying each group’s adoption rate. When risk-averse people make up a smaller fraction of the population than risk-taking people, then it would be more beneficial in reducing epidemic size to have the adoption rates of the two groups be more similar (i.e. more homogeneous) as that would imply risk-taking people (which are then the majority of the population) would have a similar adoption rate to risk-averse people. As an increasingly larger fraction of the population is composed of risk-averse people, then it becomes increasingly beneficial in reducing epidemic size to have the adoption rates of the two groups be more different (i.e. more heterogeneous) as the deleterious effects of highly risk-taking people (which are then the minority of the population) can be mitigated by the large presence of risk-averse people.

### Epidemic Size Does Not Necessarily Decrease with an Increasing Number of Risk-Averse People

General intuition would suggest that as one increases the number of risk-averse people in the population that the overall epidemic size would go down. However, the introduction of the adaptive behavior mechanism allows for regimes where this is no longer strictly the case. Thus, it is no longer a guarantee that decreasing the population’s overall risk tolerance will always improve epidemic outcomes.

Consider Figure 6a, where we find a small region after the transition to the underdamped regime where there is an increase in the epidemic size when increasing the fraction of the population that are risk-averse.

**Figure 6:**
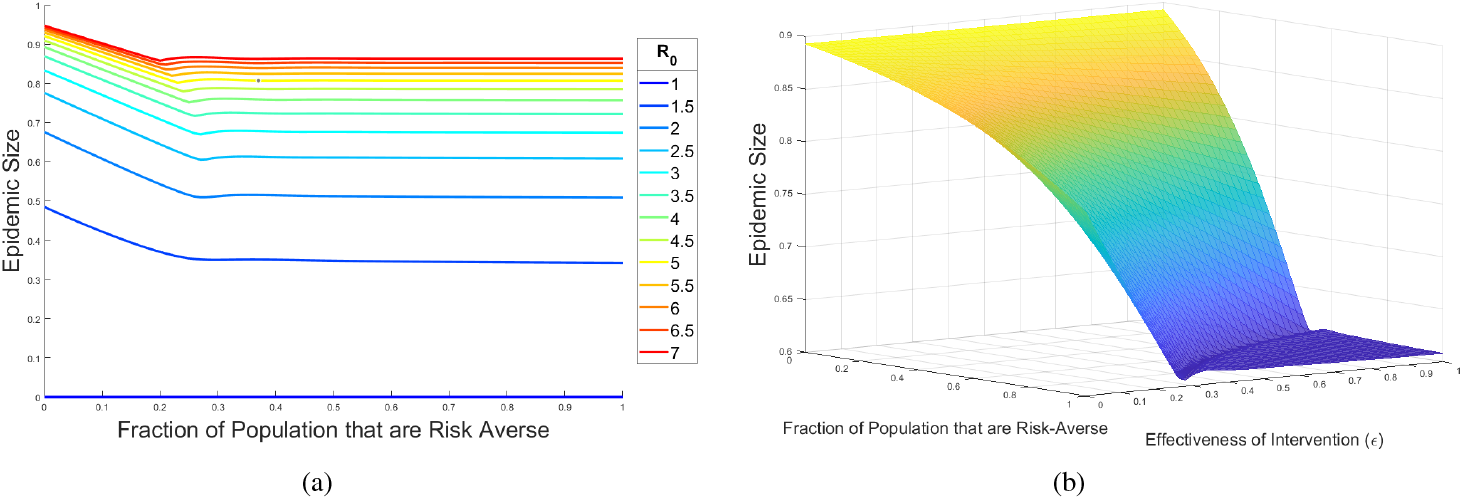
(a) Final epidemic size as a function of the proportion of the population that are risk averse (*S*_1_). Parameter values: *λ*_1_ = 10, *λ*_2_ = 0.5, *ϵ* = 1, *δ*_1_ = *δ*_2_ = 0.1. Initial conditions: *I*(0) = 10^*−*7^, *P*_1_(0) = *P*_2_(0) = 0, *R*(0) = 0. (b) Same as in (a) except now the effectiveness of the intervention (*ϵ*) is allowed to vary.

If we also vary the effectiveness of the intervention as an additional axis (Figure 6b), we observe that there is a small trench in the threshold region surrounding the plateau area. This double descent suggests that the landscape can potentially be quite complicated when risk tolerance in the population is partitioned into even more groups. While the mechanism and intuition underlying this double descent remains to be determined, it is possible that it is caused by peculiar transient dynamics that are occurring at the beginning of outbreaks in that parameter regime [54].

## 3 Discussion and Conclusions

In this paper, we have proposed a simple model to model heterogeneity in risk tolerance levels in the population. We find that including a behavioral mechanism for adopting interventions that adapts with the level of infections greatly expands the variety in epidemic dynamics and outcomes that can occur.

The general picture from the findings suggest that epidemic dynamics under adaptive intervention adoption fall into either an underdamped regime or an overdamped regime. The underdamped regime has a special property in which the epidemic size equals the herd immunity threshold exactly, which means no epidemic overshoot occurs. The system can be driven into this regime when protection mechanisms (such as numbers of risk-averse people, intervention effectiveness, and duration of intervention usage) are increased to a sufficiently high level. This regime is also marked by damped oscillations in the phase space of infecteds and susceptibles. In direct contrast, the overdamped regime closely resembles the dynamics of a simple *SIR* model without behavior, in which there are no oscillations and a non-zero overshoot, which makes the epidemic size greater than the herd immunity threshold. While we have limited our numerical exploration of the model to two groups of different risk-tolerance levels, in principle our framework can accommodate an arbitrary number of risk-tolerance groups. While the combinatorial increase in parameter space make it difficult to thoroughly explore larger models, we might naively expect there to be a complex landscape of outcomes, which might reveal other counter-intuitive phenomenology akin to the non-monotonic dependence of epidemic size with increasing number of risk-averse people for two groups.

We have looked for some evidence in the historical data on outbreaks for these damped oscillations due to cycles in adoption and relaxation of interventions. While such data is in very limited supply, previous analysis suggested that relaxation of social distancing measures may have led to multiple waves of infection in the Spanish flu of the early 20th century [55, 56]. Dating back to the time of the bubonic plague, there is data from an outbreak in 1636 in the parish of St. Martin in the Fields, which showed how relaxation of quarantining and isolation measures lead to a smaller secondary wave of infections [57]. More recently, premature relaxation of interventions during the COVID pandemic might have led to secondary outbreaks [58]. The complex milieu of disease, biological, behavioral, political, economic, and cultural factors however make it is difficult to isolate cause-and-effect in real-world epidemics [59].

The results here are reminiscent of feedback control systems commonly studied in control theory. Here the set point is the herd immunity threshold, which is determined by the basic reproduction number (*R*_0_). The ability for the population to reach this set point for epidemic size without additional overshoot depends on the effectiveness of the feedback mechanism from coupling intervention usage to the number of infected people. In the model presented, the adoption of interventions is a continuous process, in which the different groups are constantly reacting to the level of infections without requiring any notion of time or thresholds. In contrast, existing research on mitigation have considered more active control where activation and intervention timing play a key role [60–62]. Future work may explore how to synergistically utilize both active and continuous mechanisms for control.

The inclusion of heterogeneity in risk tolerance and adaptive adoption of interventions leads to several unexpected conclusions. We find that increasing heterogeneity in risk tolerance levels in the population can lead to either an increase or decrease in the epidemic size. The direction of the trend depends nonlinearly on the composition of the population in terms of the ratio of risk-averse to risk-taking individuals and their respective intervention adoption rates. This adds to a small literature that demonstrates how heterogeneity can actually lead to a larger epidemic [27, 63].

Interestingly, these results on heterogeneity also can be used to address the question of whether distributed or centralized control of mitigation results in a smaller outbreak. Control of factors such as mobility may be more practically achieved in a more centralized and unified fashion [64, 65], whereas a distributed approach may be more appropriate when considering factors such as speed and individual agency. In a centralized scenario, a single entity controls the dynamics. That situation has an exact correspondence to the homogeneous population considered here, where all individuals respond in unison. Distributed control allows for more localized control, such as individuals or small groups deciding if they want to mask or social distance. This corresponds to the heterogeneous scenarios considered here, where there are multiple groups each with differing risk tolerance level. The results suggest that in some scenarios a single, coordinated response would be better for mitigation, whereas in other parameter regimes, a more decentralized strategy would be more optimal.

We also find that increasing overall protection mechanisms does not always result in a monotonic decrease in epidemic size. In scenarios when the adoption rate begins to approach the transmission rate, near the critically damped boundary a nonmonotonicity can arise. This suggests that when intervention usage and effectiveness are tenuous, the dynamics become more complex and predicting what epidemic outcomes will result becomes significantly more difficult. Understanding how these nonlinear effects combine with other biological and behavioral heterogeneities will be important to explore in future work.

## 4 Methods

### Defining the *λ* Homogeneity Index

The *λ* homogeneity index is defined as follows. We will assume the initial condition that at the beginning of the dynamics, the total population is composed of the fraction of the population in the low risk tolerance group *x*_1_, the high risk tolerance group *x*_2_, or the infected class. We will assume the fraction of the population initially infected is sufficiently small so that size of the two susceptible compartments is given by *x*_1_ and 1 − *x*_1_ respectively.

Define a homogeneity index parameter (*c*) that captures the degree of homogeneity between the two adoption rates, *λ*_1_ and *λ*_2_. We define the parameter domain to be the unit interval, *c* ∈ (0, 1]. An index value of *c* = 1 indicates *λ*_1_ = *λ*_2_, while decreasing the index value towards 0 increases the difference between *λ*_1_ and *λ*_2_.

We can define the average adoption rate as being either a geometric or arithmetic mean of the two adoption rates. The choice one makes is arbitrary, so we present prescriptions for both routes. In both cases, we will map the level of homogeneity to the unit interval.

### Geometric Average

Let the geometric average of the two adoption rates be given by *λ*_Geometric Average_.

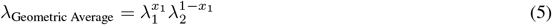

Define *λ*_2_ as a fraction between 0 and 1 of the average *λ*.

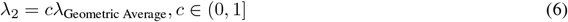

These two equations combine to define *λ*_1_.

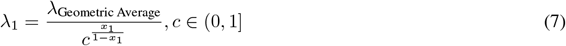

### Arithmetic Average

Let the arithmetic average of the two adoption rates be given by *λ*_Arithmetic Average_.

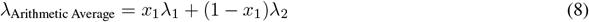

Define *λ*_2_ as a fraction between 0 and 1 of the average *λ*.

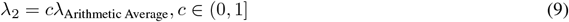

These two equations combine to define *λ*_1_.

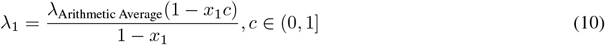

Again, an index value of 1 indicates *λ*_1_ = *λ*_2_, while decreasing the index value towards 0 increases the difference between *λ*_1_ and *λ*_2_.

### Homogeneity Index in Larger Groups

The above parameterization of homogeneity is useful for in the setting of a two-group model because the entirety of the possible difference in heterogeneity between *λ*_1_ and *λ*_2_ can been mapped to the unit interval. This allows us to more easily see the qualitative shift in trend in phenomenology that prompted this section of text in the first place. An alternative parameterization might have used the variance between the two adoption rates. However, the domain of such a parameterization is unbounded, making it impossible to explore fully numerically. Moving beyond two groups would necessitate a different parameterization (such as through using a variance or coefficient-of-variation approach).

## Author Contributions

M.M.N., B.E.C., C.M.S.-R., B.T.G., and S.A.L. designed the study. M.M.N., A.S.F., M.A.C. performed the simulations and calculations. M.M.N. wrote the manuscript. All authors contributed to interpreting the results and editing the manuscript.

## Acknowledgments

The ideas and conversations that sparked this work originated during a special session on the Mathematics of Infectious Diseases: A Session in Memory of Dr. Abdul-Aziz Yakubu, III at the 2024 Spring Sectional Meeting of the American Mathematical Society. The authors would like to thank the organizers of that session: Abba Gumel, Daniel Cooney, and Chadi Saad-Roy.

## Data Availability

The code used to generate all Figures is provided in the Supplemental Material.

## Supplemental Material

### Analysis of the Model When Individuals Respond to the Incidence Rate

The rate at which intervention adoption occurs may be driven by individuals considering information such as the epidemic incidence rate (e.g. cases per day), the total number of infected individuals in the population (e.g. total number of active cases), and mortality rate (e.g. deaths per day) [22]. Here we will consider the first case, where individuals adopt interventions based on the incidence rate for infections. Recall that the incidence rate is given by 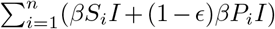. Parameterizing each person’s individual risk tolerance by *λ*_*i*_, let us assume each individual adopts an intervention at rate 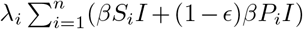. Then, if there are *S*_*i*_ number of people that behave exactly the same (i.e. have the same level of risk-aversion), then at the population scale there is a collective adoption rate of 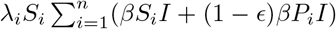. The same reasoning holds for each of the *n* tolerance levels. The corresponding equations for this model are given by(11-14).

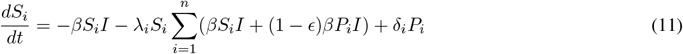

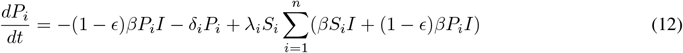

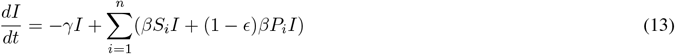

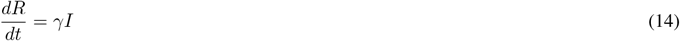

When comparing the final epidemic size and epidemic trajectories between this model and the model in the main text where individuals adopt interventions at a rate that is based on the total number of infected people (Figures S4-S5), the results are indistinguishable (Figures S1-S2).

**Figure S1:**
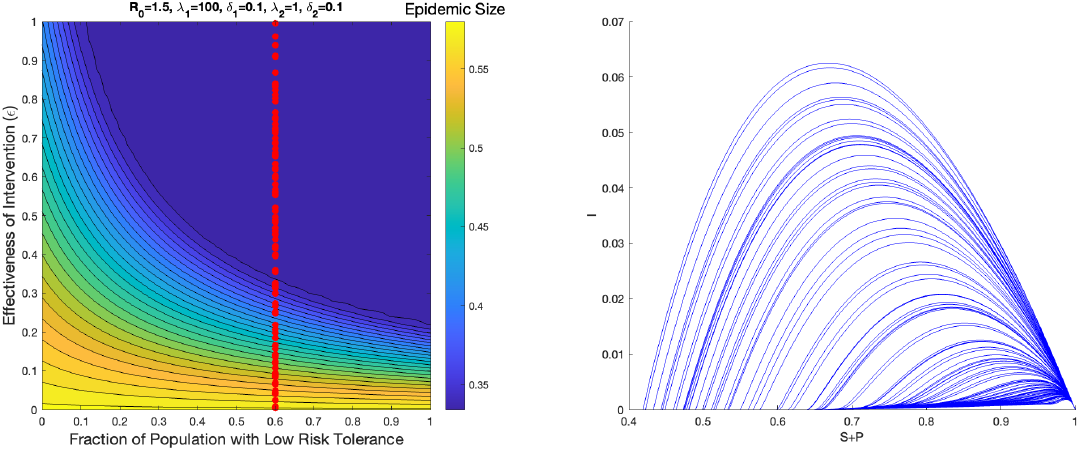
Left. Epidemic size as a function of varying the fraction of the population that are low-risk tolerance (i.e. those with higher *λ*) when individuals react to incidence rate. Right. Corresponding orbits in the I versus S+P plane for the sampled points in parameter space when the fraction of the population that are low-risk tolerance has been fixed. Parameter values: *β* = 1.5, *γ* = 1, *λ*_1_ = 100, *δ*_1_ = 0.1, *λ*_2_ = 1, *δ*_2_ = 0.1,. Initial conditions: *I*(0) = 10^*−*^8, *P*_1_(0) = *P*_2_(0) = *R*(0) = 0.

Thus, given the equivalence in results, we have gone with the mathematically simpler and cleaner model based on total number of infected people in the main text.

**Figure S2:**
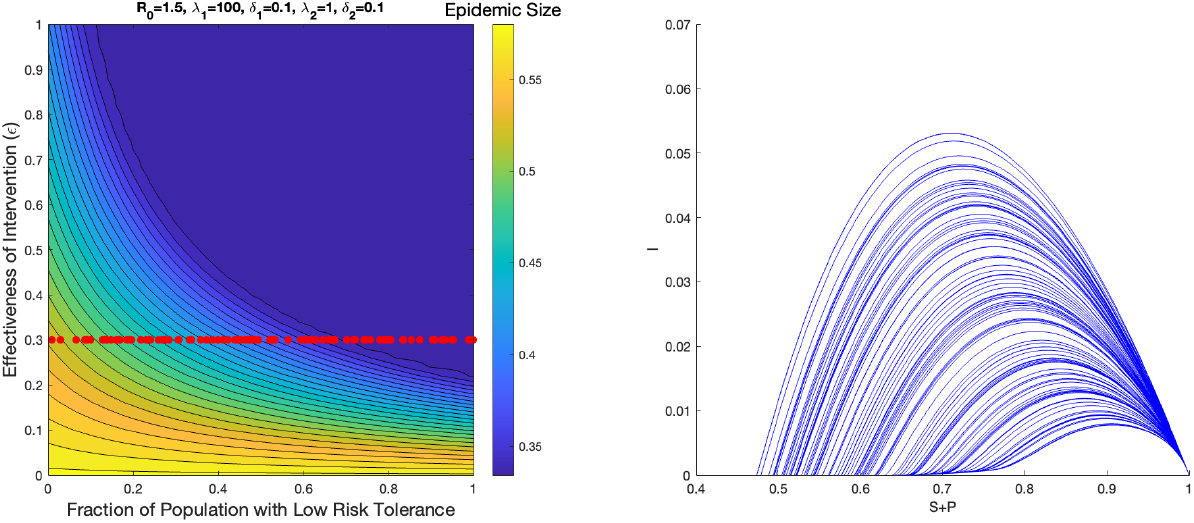
Left. Epidemic size as a function of varying the fraction of the population that are low-risk tolerance (i.e. those with higher *λ*) when individuals react to incidence rate. Right. Corresponding orbits in the I versus S+P plane for the sampled points in parameter space when the intervention effectiveness has been fixed. Parameter values: *β* = 1.5, *γ* = 1, *λ*_1_ = 100, *δ*_1_ = 0.1, *λ*_2_ = 1, *δ*_2_ = 0.1,. Initial conditions: *I*(0) = 10^*−*^8, *P*_1_(0) = *P*_2_(0) = *R*(0) = 0.

### The Herd Immunity Threshold is Set by a Complex Interplay Between Transmission (*R*_0_), Behavior, and Intervention Effectiveness (*ϵ*)

While we could make some analytical calculations for the epidemic size in the homogeneous model, the heterogeneous two group case requires a numerical approach to find the plateau region. In general, it is set by a highly nonlinear interaction between the transmission (*R*_0_), behavior as determined by the fraction of the population that are risk-averse and risk-taking, and the effectiveness of the intervention (*ϵ*).

Consider the following progression of Figures (Figure S3), where in each subsequent Figure, the effectiveness of the intervention in blocking transmission (*ϵ*) is increasing.

**Figure S3:**
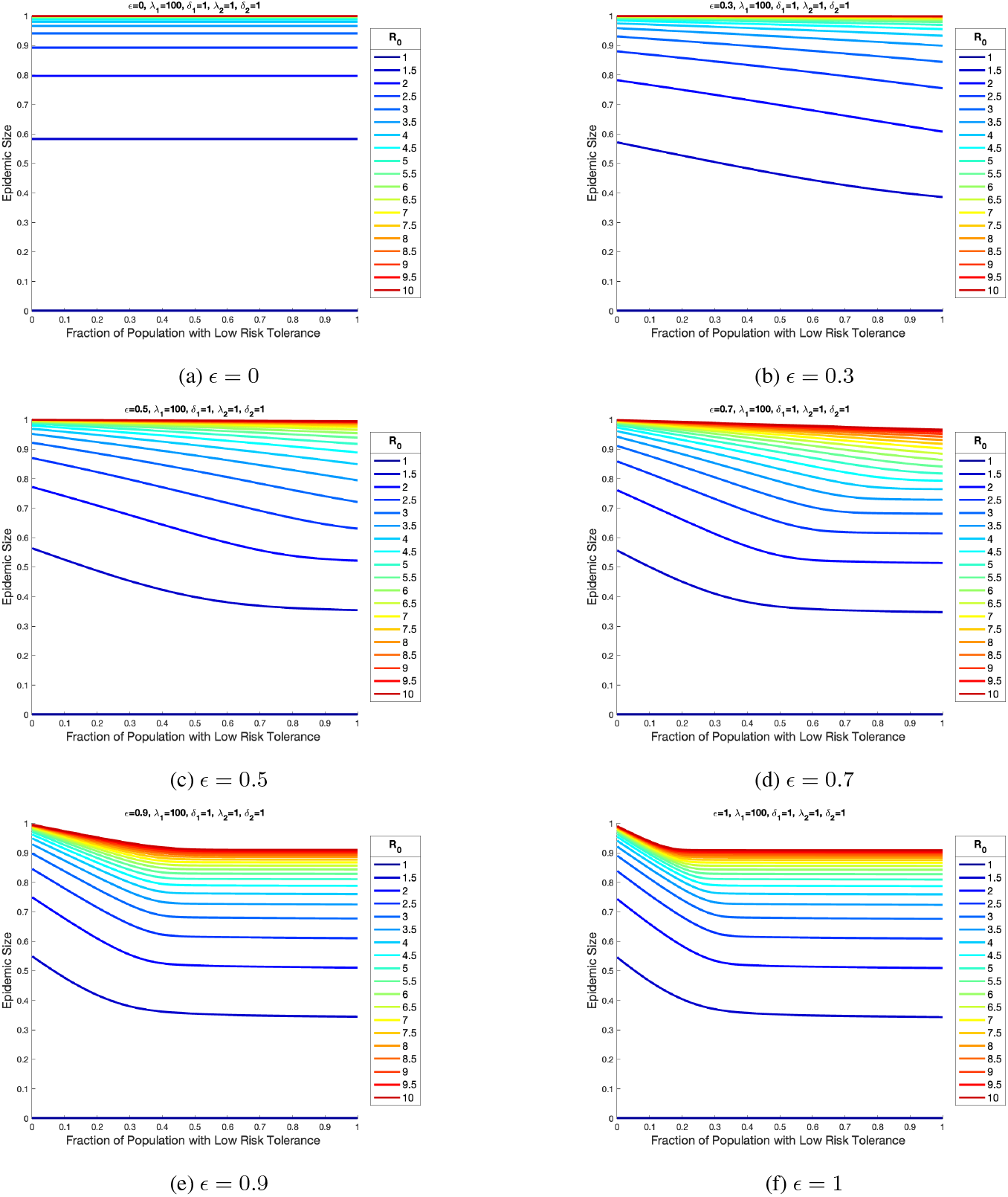
Final epidemic size versus fraction of population that are risk-averse (*S*_1_) with a progressive increase in intervention effectiveness (*ϵ*). Parameter values: *λ*_1_ = 100, *δ*_1_ = 1, *λ*_2_ = 1, *δ*_2_ = 1. Initial conditions: *I*(0) = 10^*−*7^, *P*_1_(0) = *P*_2_(0) = 0, *R*(0) = 0.

### Comparing Orbits Inside and Outside of the Plateau Region of Herd Immunity

The following Figures sample more orbits for the Figure considered in the main text.

**Figure S4:**
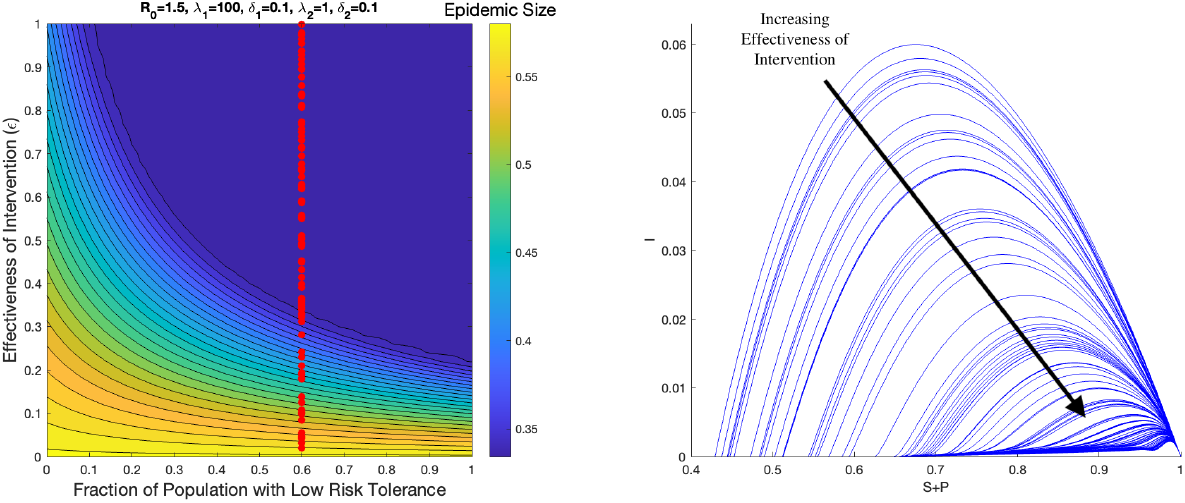
Left. Epidemic size as a function of varying the fraction of the population that are low-risk tolerance (i.e. those with higher *λ*). Right. Corresponding orbits in the I versus S+P plane for the sampled points in parameter space when the fraction of the population that are low-risk tolerance has been fixed. Parameter values: *β* = 1.5, *γ* = 1, *λ*_1_ = 100, *δ*_1_ = 0.1, *λ*_2_ = 1, *δ*_2_ = 0.1,. Initial conditions: *I*(0) = 10^*−*^8, *P*_1_(0) = *P*_2_(0) = *R*(0) = 0.

**Figure S5:**
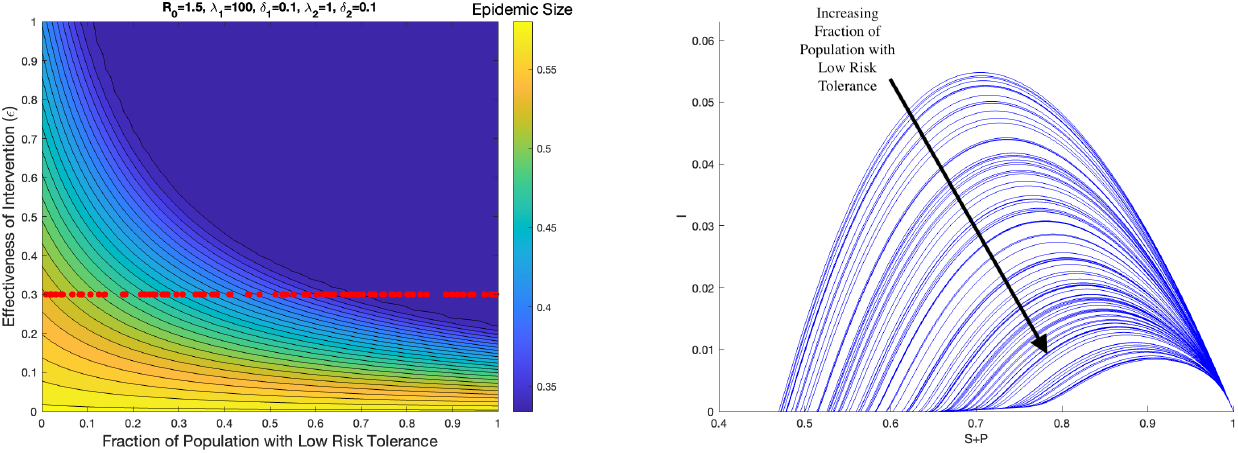
Left. Epidemic size as a function of varying the fraction of the population that are low-risk tolerance (i.e. those with higher *λ*). Right. Corresponding orbits in the I versus S+P plane for the sampled points in parameter space when the intervention effectiveness has been fixed. Parameter values: *β* = 1.5, *γ* = 1, *λ*_1_ = 100, *δ*_1_ = 0.1, *λ*_2_ = 1, *δ*_2_ = 0.1,. Initial conditions: *I*(0) = 10^*−*^8, *P*_1_(0) = *P*_2_(0) = *R*(0) = 0.

### Proving Underdamped Regime Eliminates Epidemic Overshoot

In general, the nonlinear feedback between the protected classes and infected individuals make it difficult to make analytical calculations in the full model. Under some simplifications however, we can make some progress. In this section, we derive the epidemic size at which the protection saturates in a restricted case of the homogeneous model (*n* = 1).

Let us consider the homogeneous model in the limit of an intervention with perfect effectiveness (i.e. *ϵ* = 1). We will consider the case where the recovery rate from infection and relaxation rate for interventions are comparable (i.e. *γ* = *δ*). Since the equation for recovered individuals can be ignored since the population is closed (*S* + *P* + *I* + *R* = 1), this reduces the dynamics to the following system:

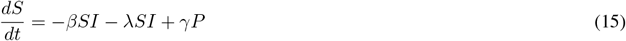

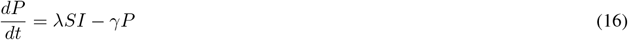

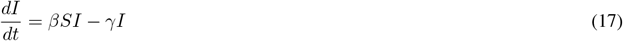

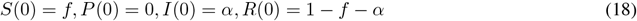

The initial conditions sets the number of individuals initially susceptible to be *f*, the number of initially infected is assumed to be small *I*(0) *<<* 1, and the remainder of the population is already immune to infection (i.e. recovered). To ensure that the epidemic initially grows in size, we assume that 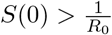. This follows from comparing the incidence term (*βSI*) to the recovery term (*γI*) in17.

To find the asymptotic behavior for *S*, we will attempt to eliminate *P* from(15). We start first by seeking an equation that relates the *P* and *I* compartments. Consider the following ansatz that considers the difference between the two compartments:

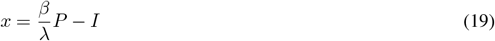

Differentiating this equation with respect to time and using(16)-(17) yields:

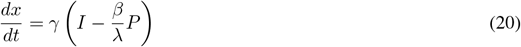

Using(19), this simplifies to 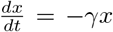, which has a critical point at *x* = 0. At *x* = 0, we obtain that 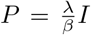, indicating a regime where the behavior of *P* and *I* scale linearly with each other. Combining this with(15) yields:

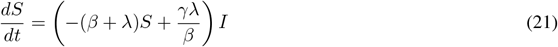

Now that we have an equation that is linear in *I*, we are in a good position to find a final size relationship for the number of susceptibles. To start we take the ratio of(17) and(21).

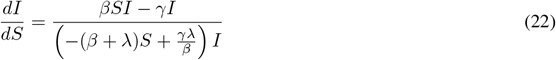

Using the partial fractions 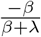 and 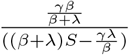, we get upon indefinite integration of(22) that 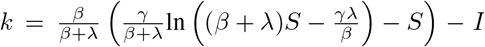, where *k* is a constant that holds throughout the trajectory of the dynamics. Thus, considering the values of *S* and *I* at the beginning of the epidemic (*t* = 0) and the end of the epidemic (*t* = ∞) and using the conditions that *I*(∞) = 0 and *I*(0) ≈ 0 yields the following transcendental equation for the final epidemic size.

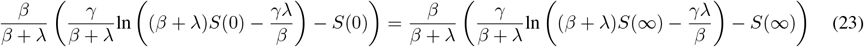

Since 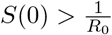, then the argument of the logarithm on the left hand side must be positive, and subsequently the left hand side evaluates to a real number. Due to the equality, the right hand side must also evaluate to a real number, implying the argument of the logarithm on the right hand side must also be positive. Positivity implies the following inequality for the lower bound for the final number of susceptibles:

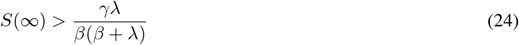

An upper bound can be given by simply noting that in the long time limit, the recovery term (*γI*) must be at least as large as the incidence term (*βS*(∞)*I*) in(17), otherwise the epidemic would still be growing. This implies 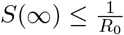.

To summarize, when the interventions are perfectly effective, the rate of relaxation from the protected class is equal to the rate of recovery from infection, and the number in the protected class scales linearly with the number of infected, then the final fraction of susceptibles is bounded as follows:

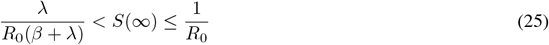

We see in the parameter limit of when the adoption rate of interventions is very fast compared to the transmission rate (i.e. *λ* >> *β*), that the lower bound reduces to 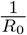. Since both bounds now coincide, then *S*_*∞*_ must equal that value. Interestingly this corresponds to the herd immunity threshold of the standard SIR model. As the overshoot is the excess number of cases beyond the herd immunity threshold, we see that in this parameter limit there is no overshoot.

This analysis for the homogeneous case also carries over to the heterogeneous case for two groups when the adoption rate between the two groups is significantly different (i.e. *λ*_1_ >> *λ*_2_). This results in a separation of time scales in which the faster adopters quickly transition to the protected state and can essentially be treated as immune over the course of the remaining epidemic over the slow adopters. This amounts to effectively reducing the dynamics to the homogeneous model considered here where *f* and 1− *f* fractions of the population in the susceptible and recovered respectively correspond to the fraction of the population in the slow adopter (*λ*_2_) and fast adopter groups (*λ*_1_).

### Heterogeneity in Risk Tolerance through Arithmetic Averaging

There are two ways for calculating the difference (heterogeneity) in adoption rates, either through geometric or arithmetic averaging. Both can be justified, and we presented the geometric formulation in the main text. We find that the arithmetic formulation gives qualitative similar results under a suitable parameter shift (Figure S6).

**Figure S6:**
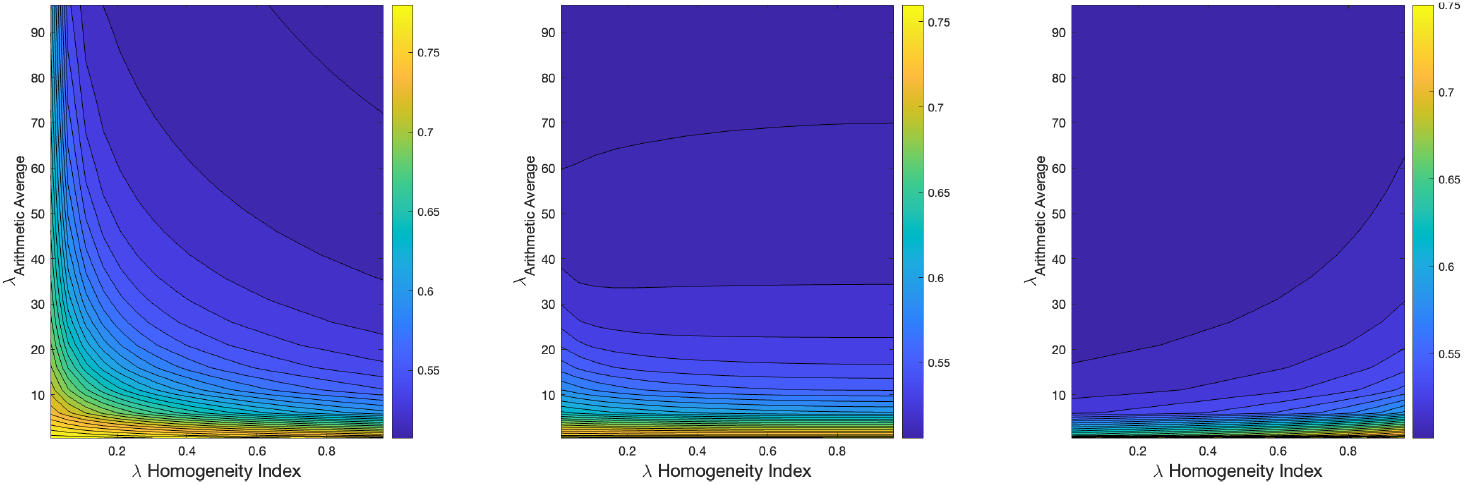
Epidemic size under differing levels of heterogeneity in the adoption rate for interventions. The mean adoption rate of the two groups (i.e. arithmetic average of *λ*_1_, *λ*_2_) is compared to the difference in the two adoption rates as parameterized by a homogeneity index (see Methods for definition). *Left* is when the fraction of the population with low risk tolerance (*x*_1_) is 0.2, *center* is when *x*_1_ = 0.5, *right* is when *x*_1_ = 0.8. Parameter values: *β* = 2, *γ* = 1, *ϵ* = 0.7, *δ*_1_ = *δ*_2_ = 0.5. Initial conditions: *I*(0) = 10^*−*^8, *P*_1_(0) = *P*_2_(0) = *R*(0) = 0.

### Heterogeneity in Relaxation Rates of Protection

In the main text we primarily assumed that the relaxation rate in both groups were homogeneous for simplicity (i.e. *δ*_1_ = *δ*_2_). However, in the real world, one might actually expect for them to be inversely correlated with the intervention adoption rates, such that if *λ*_1_ > *λ*_2_, then *δ*_1_ *< δ*_2_. The intuition is that a more risk-averse person will adopt protection faster and relax the protection at a slower rate. We numerically study if we allow for heterogeneous relaxation rates. As one might expect, our numerical results that the behavior of the epidemic size is intermediate between the two homogeneous extremes. (Figure S7).

**Figure S7:**
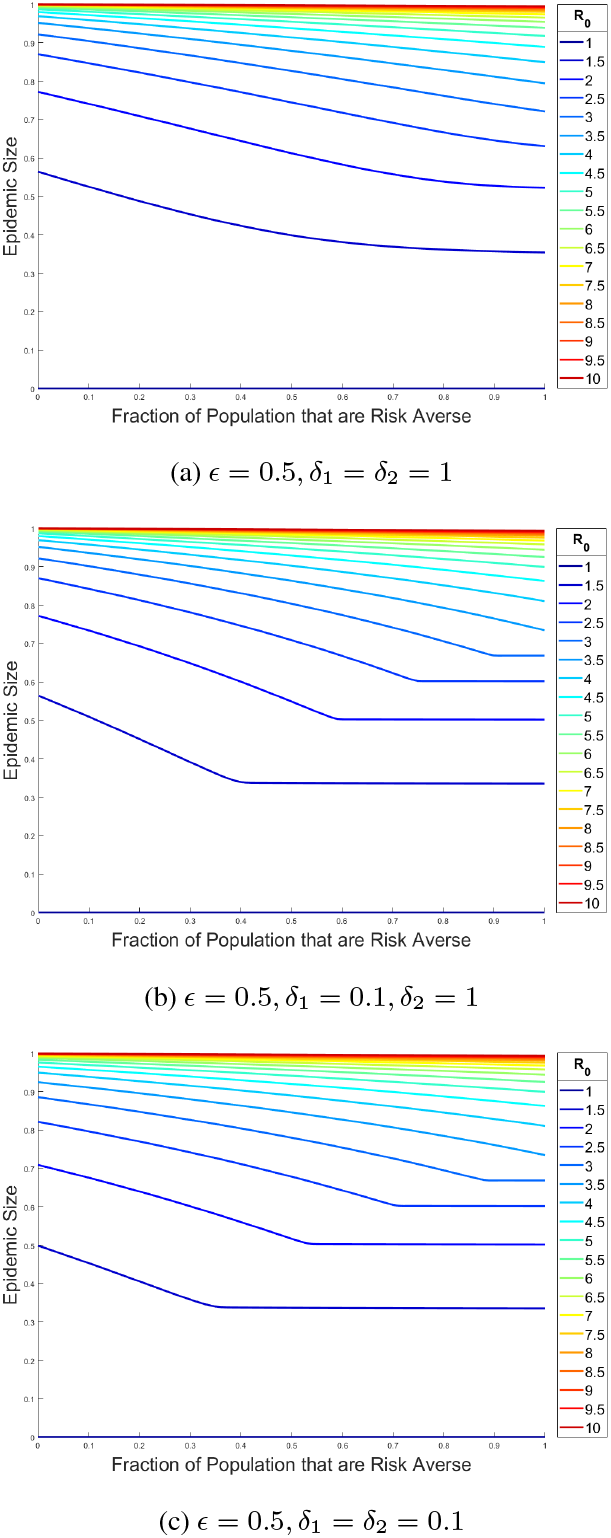
Final epidemic size versus fraction of population that are risk-averse (*S*_1_). Simulations in the left column have a higher *δ* than simulations in the right column. Parameter values: *R*_0_ = *β, γ* = 1, *λ*_1_ = 100, *λ*_2_ = 1. Initial conditions: *I*(0) = 10^*−*^8, *P*_1_(0) = *P*_2_(0) = *R*(0) = 0.

## Code to Generate Figures

Code executed in MATLAB R2023a.

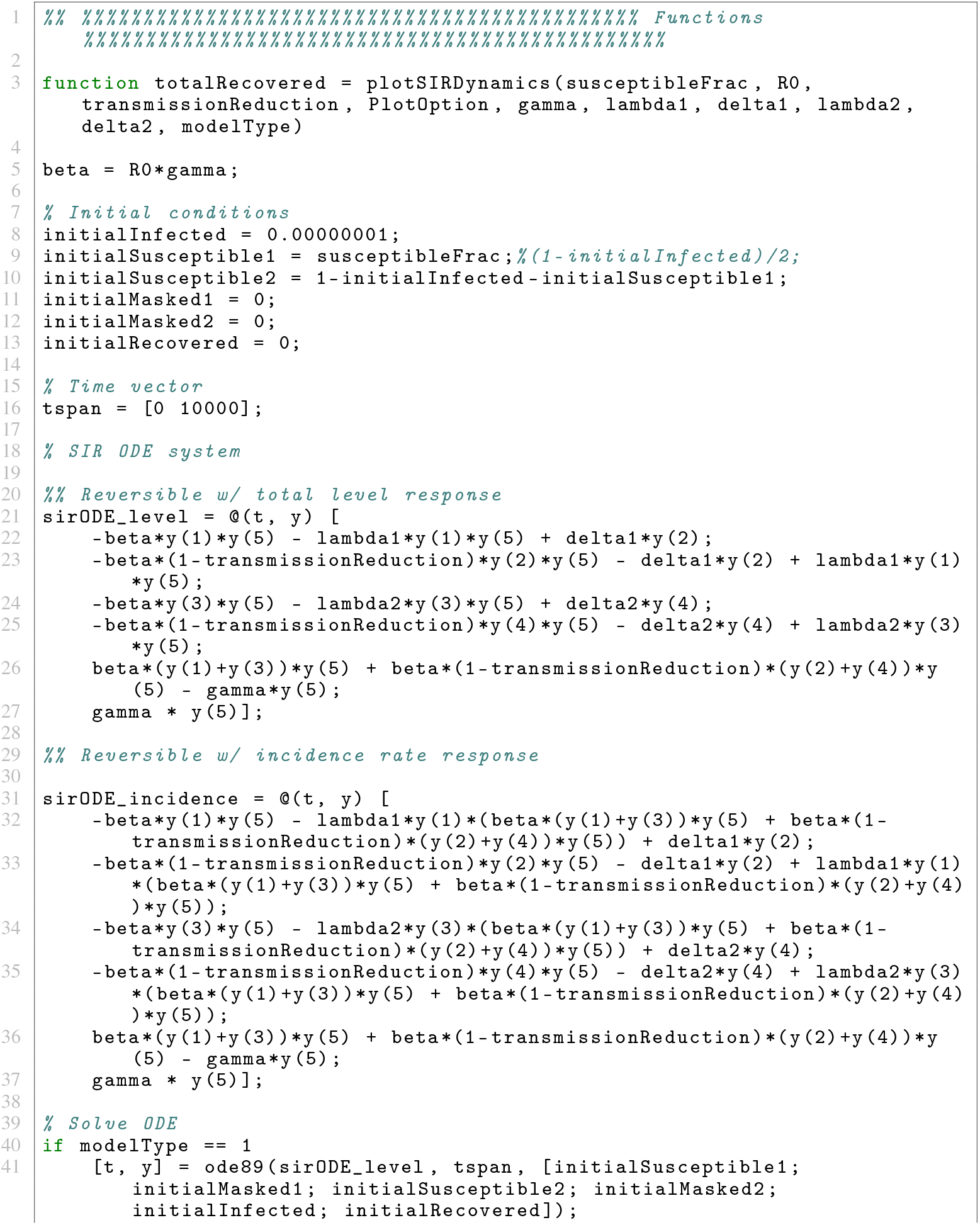
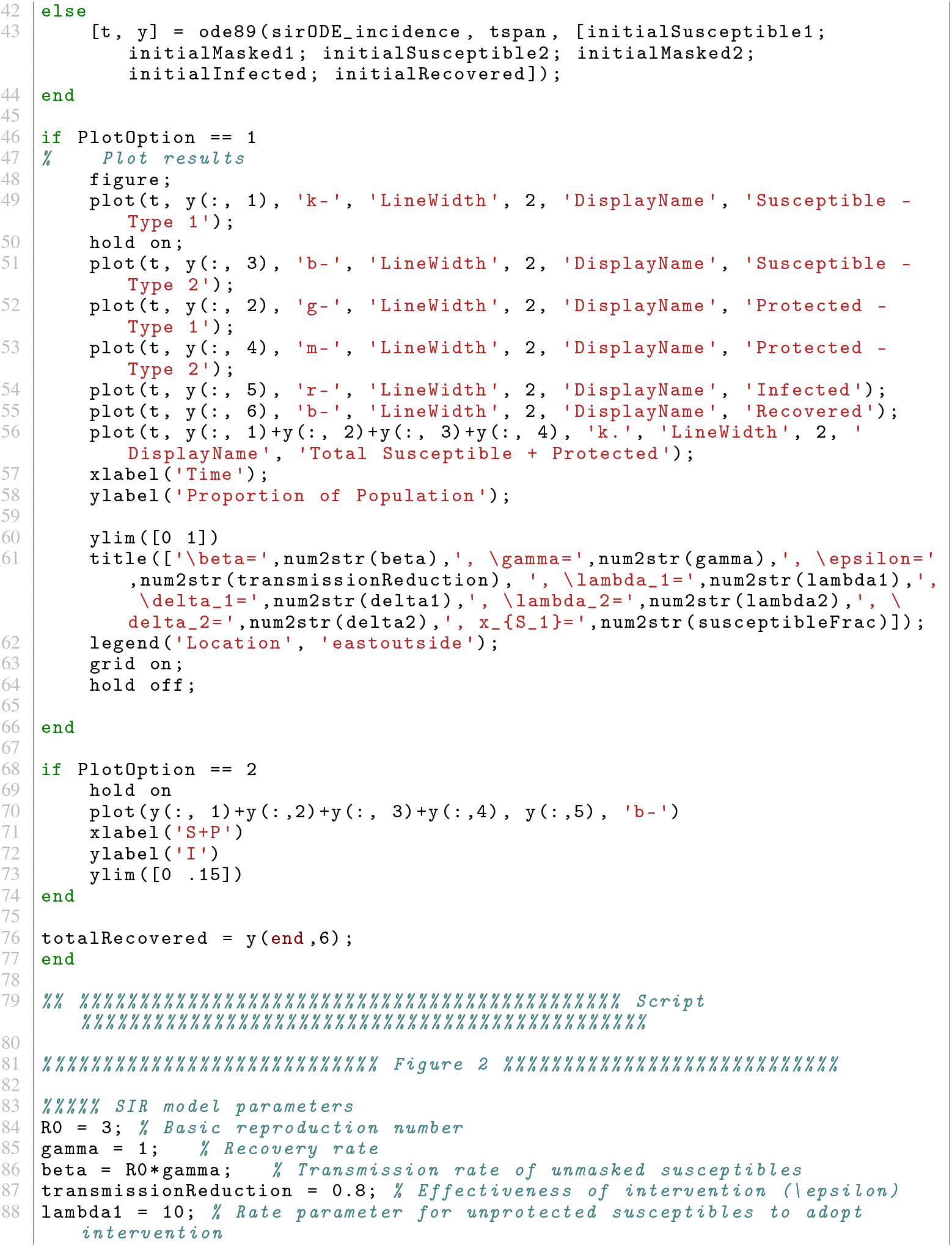
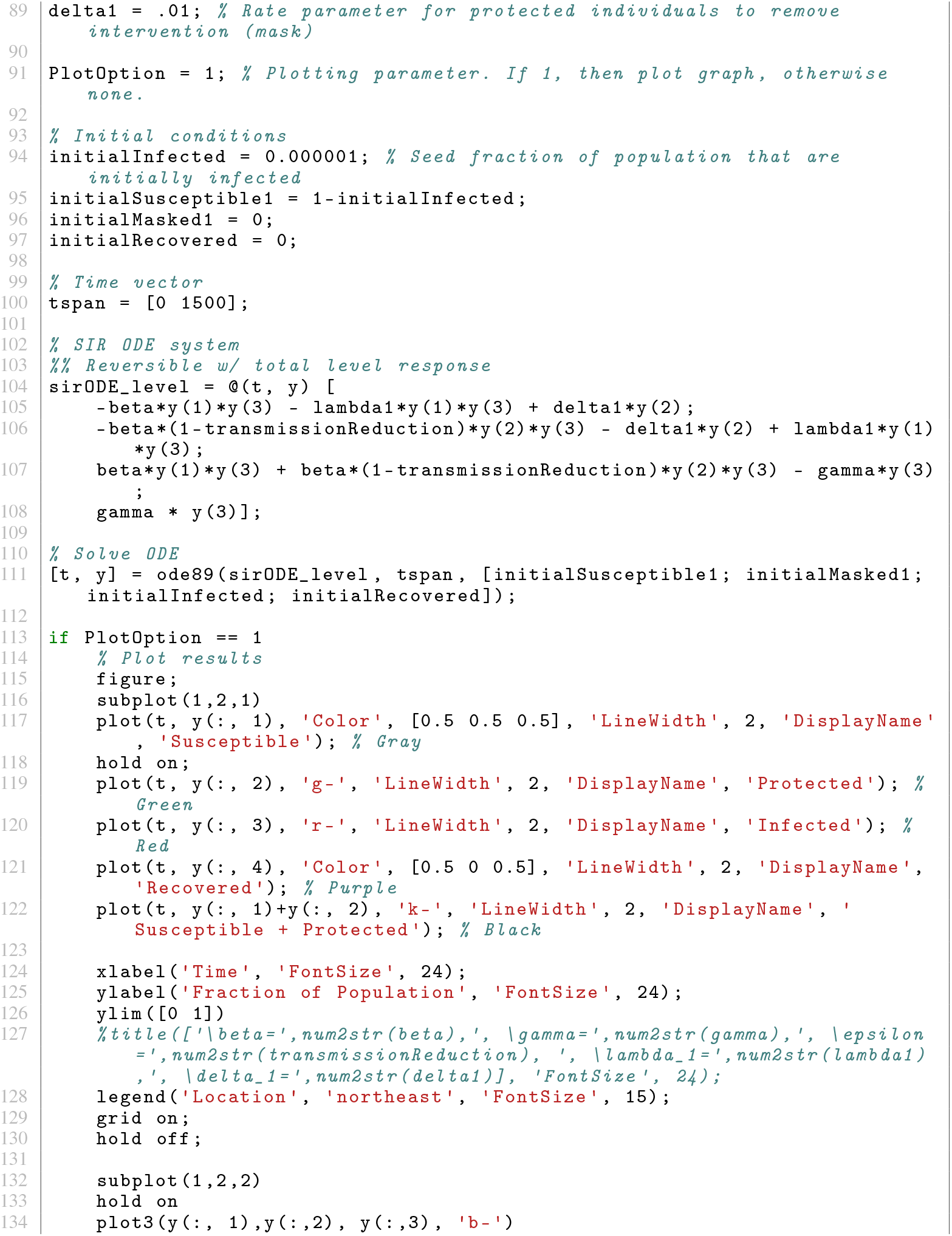
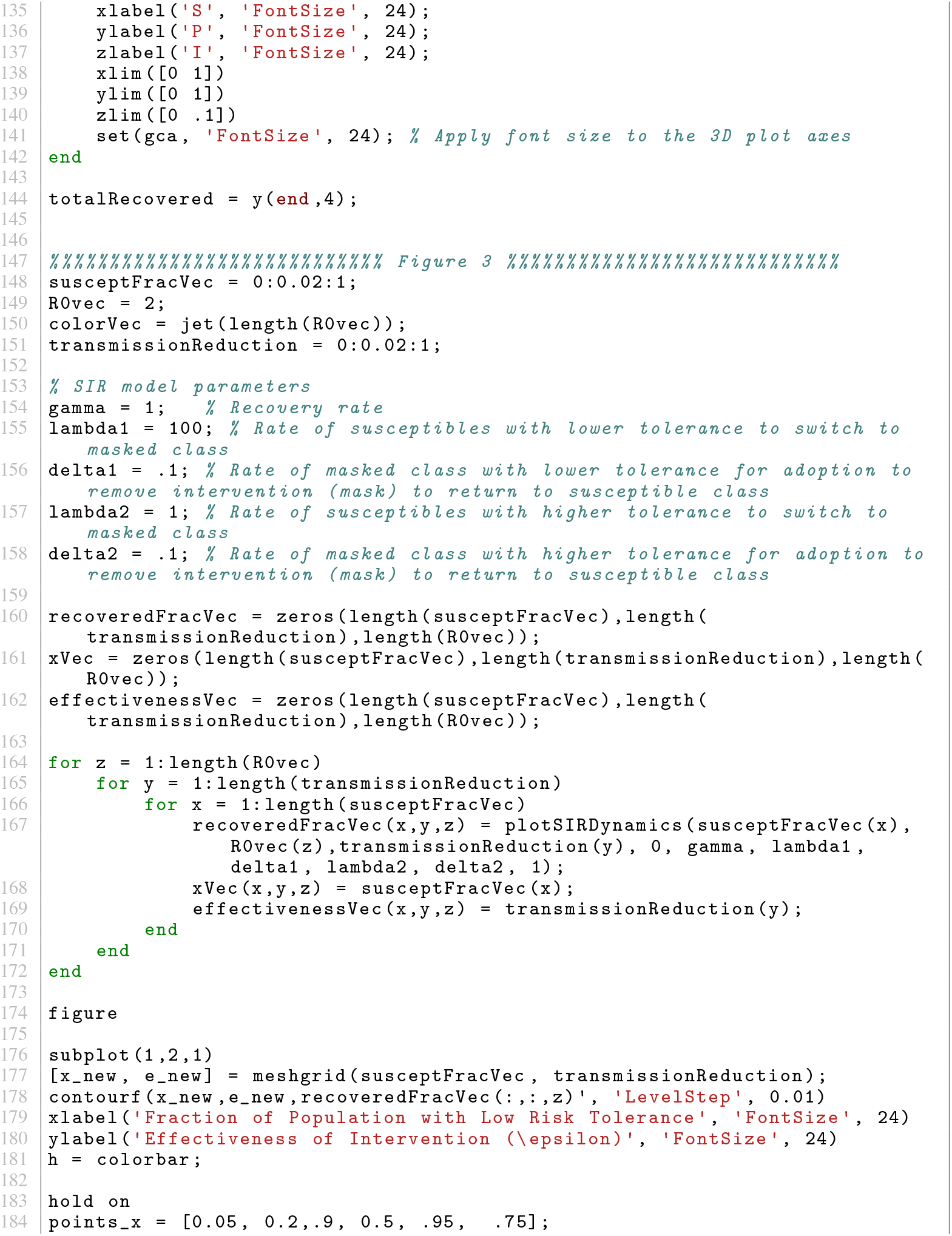
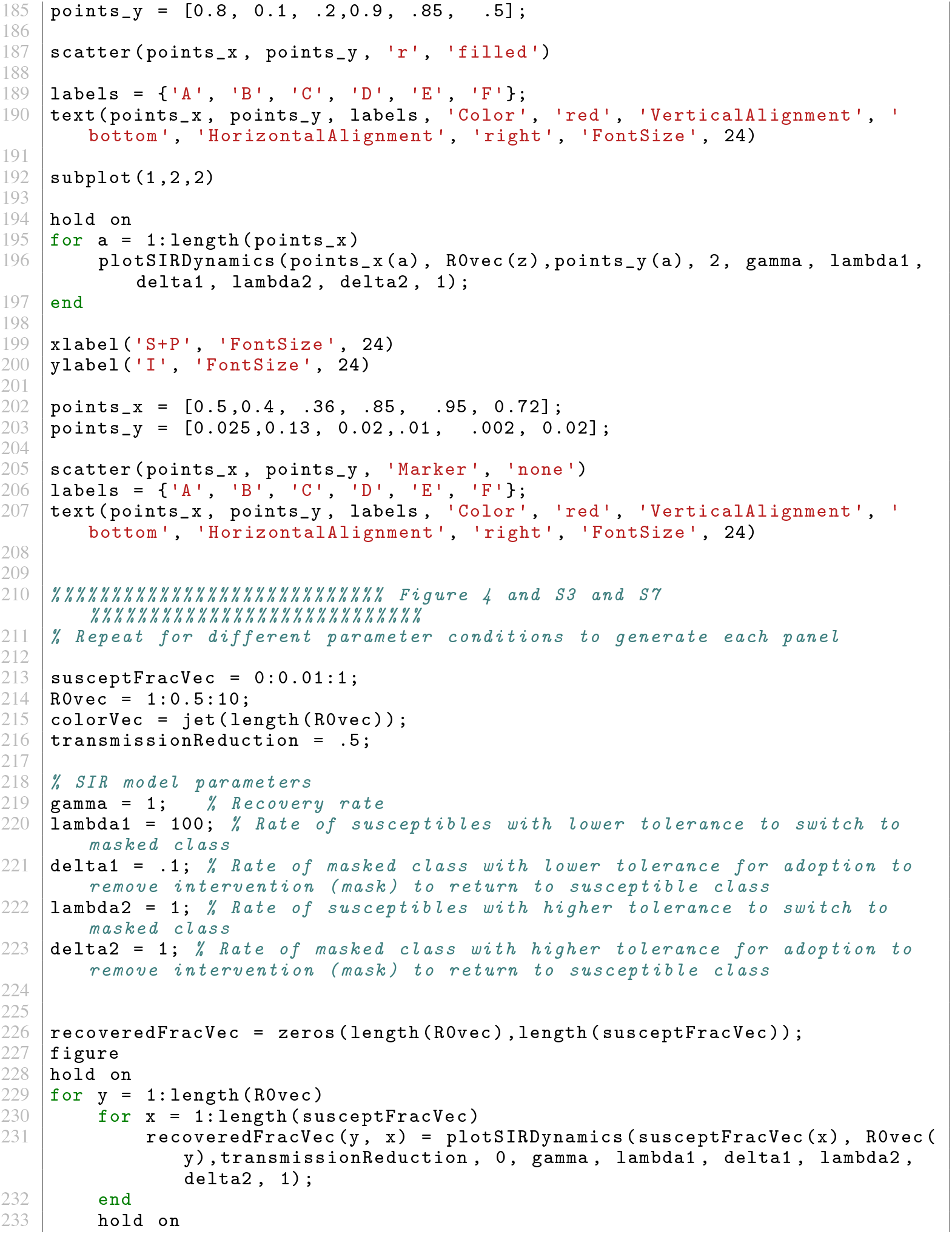
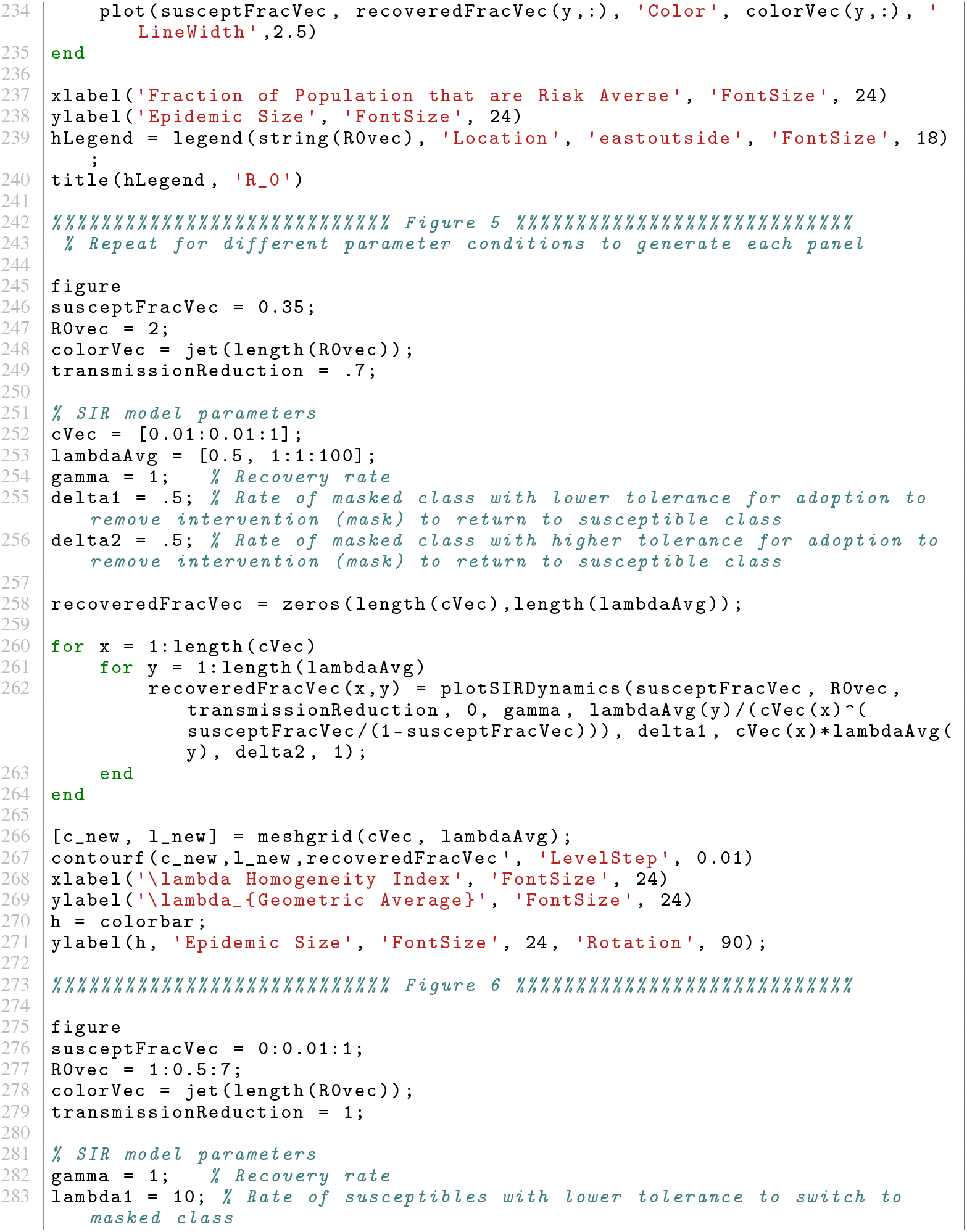
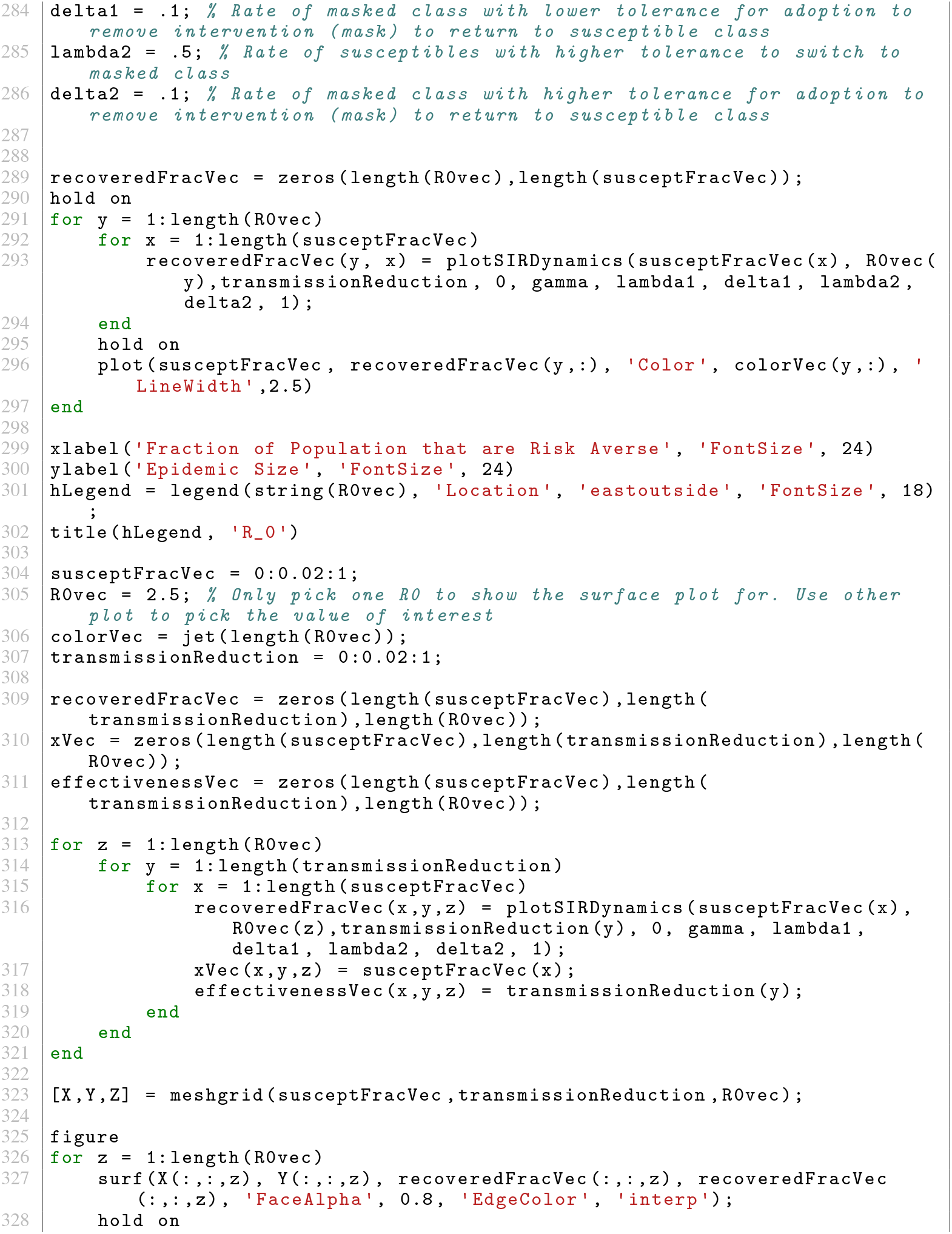
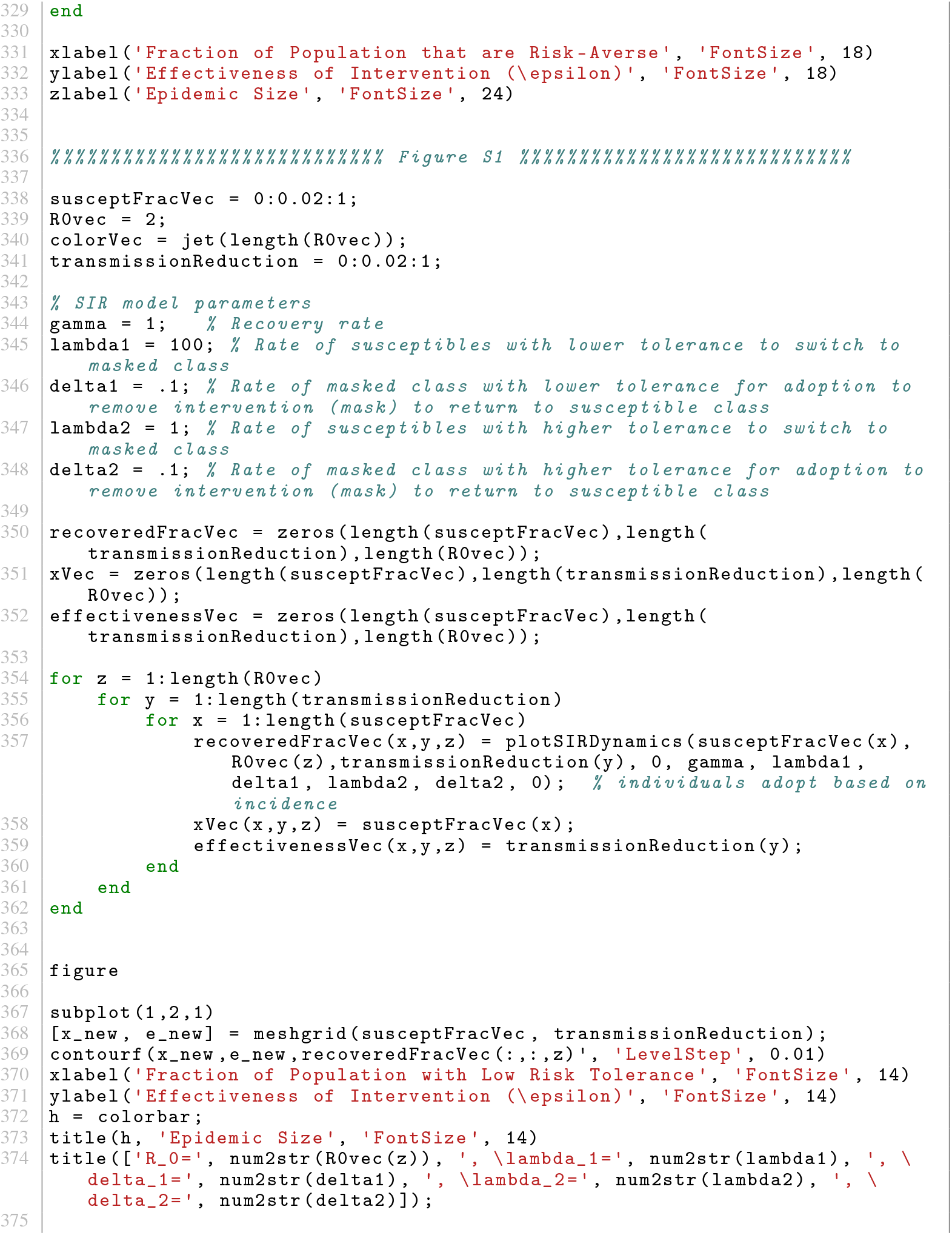
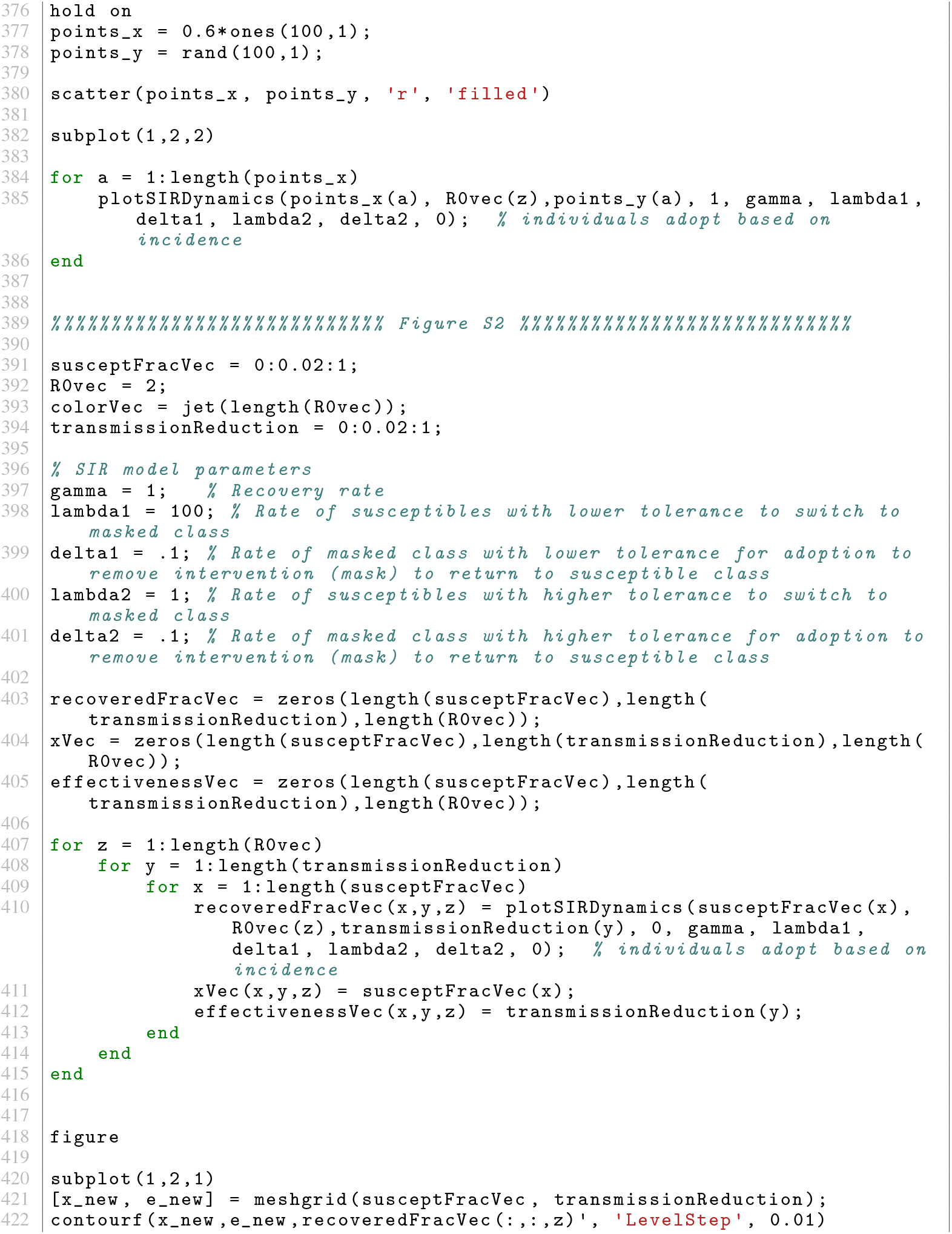
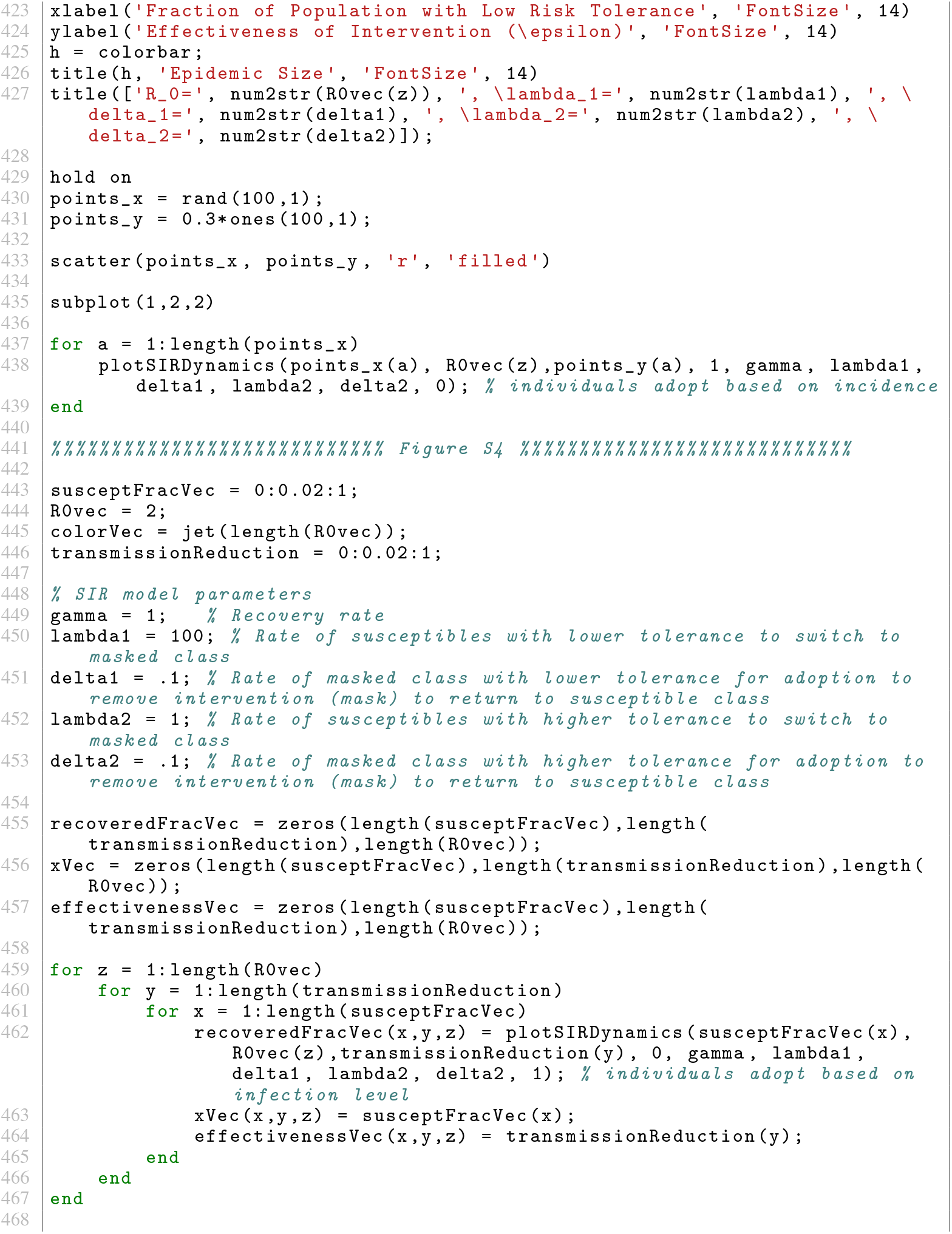
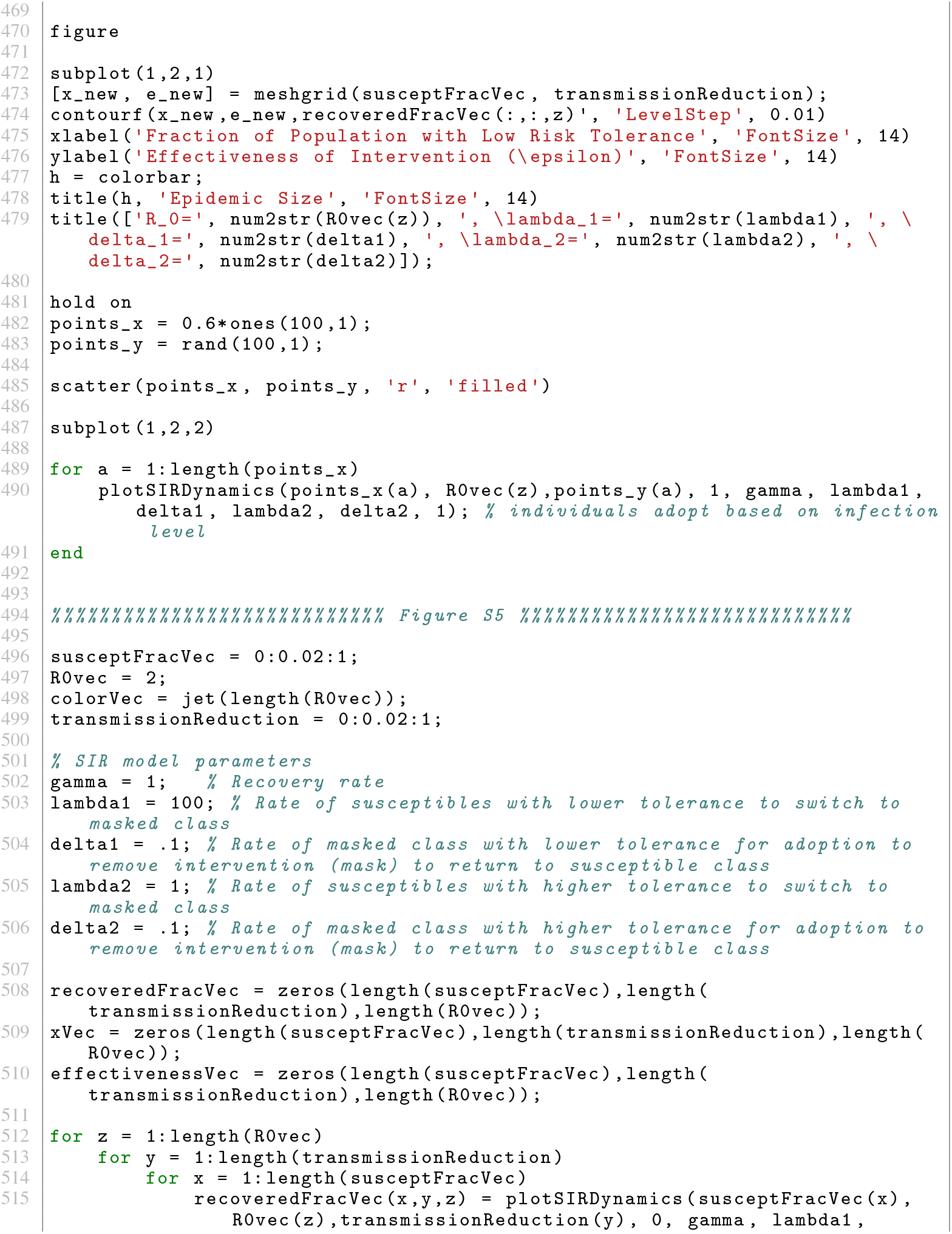
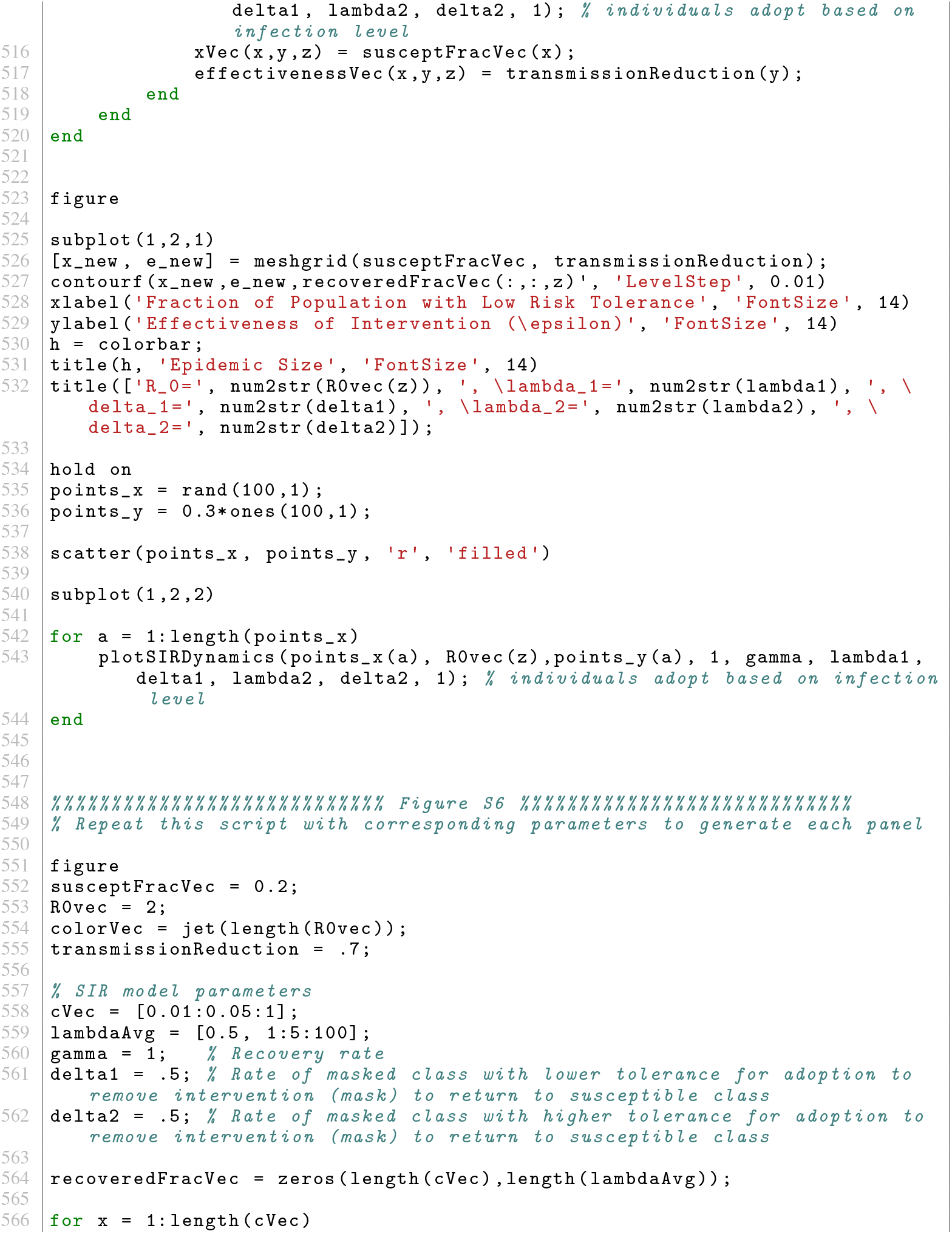
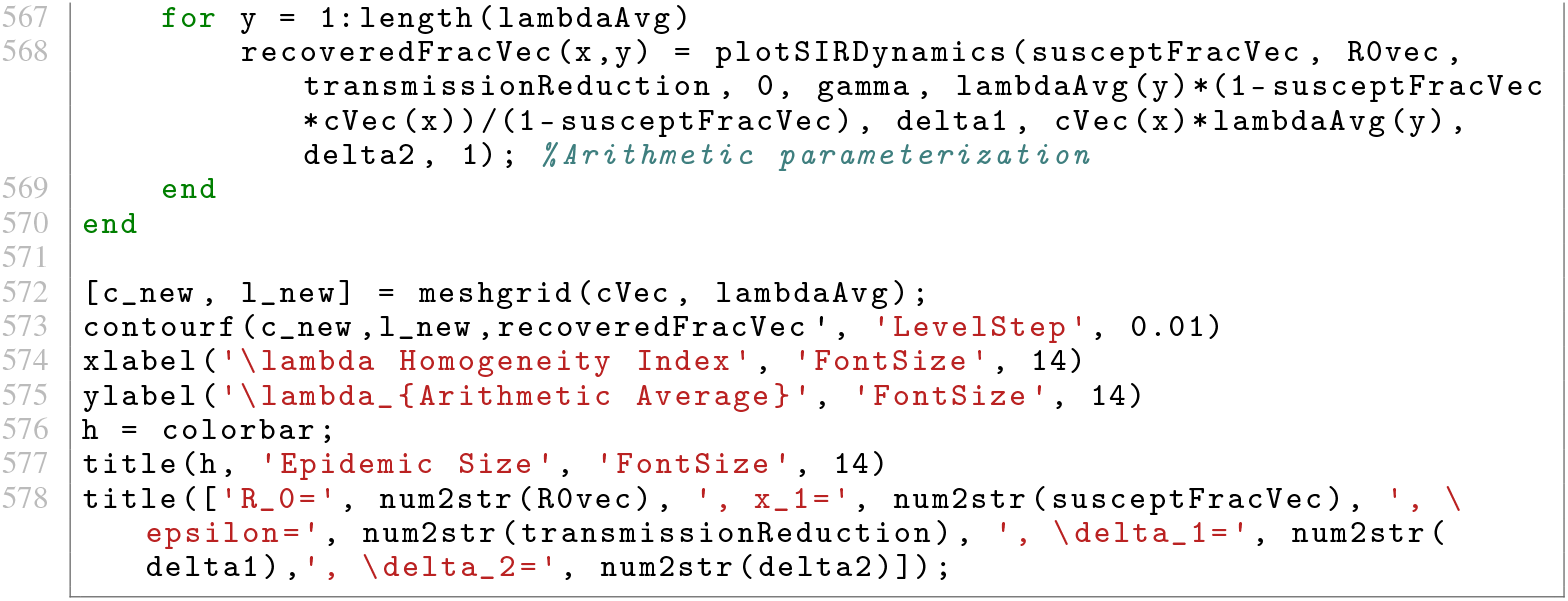

